# Genetic variation in *HIF1A* is associated with smoldering inflammation and disease progression in Multiple Sclerosis

**DOI:** 10.1101/2024.03.15.24304290

**Authors:** Antonino Giordano, Pernilla Stridh, Paolo Preziosa, Marco Pisa, Melissa Sorosina, Elisabetta Mascia, Silvia Santoro, Kaalindi Misra, Ferdinando Clarelli, Laura Ferrè, Maria Needhamsen, Ali Manouchehrinia, Miryam Cannizzaro, Thomas Moridi, Klementy Shchetynsky, Russell Ouellette, Adil Harroud, Elisabeth Sandberg, Subita Balaram Kuttikkatte, Fredrik Piehl, Lars Alfredsson, Jan Hillert, Tomas Olsson, Lars Fugger, Kate Attfield, Tobias Granberg, Maja Jagodic, Gabriele Carmine DeLuca, Mara Rocca, Massimo Filippi, Ingrid Kockum, Federica Esposito

## Abstract

**Background:** Understanding the mechanisms underlying disease progression in Multiple Sclerosis (MS) is fundamental to pave the way to treatment advances. Smoldering demyelinating inflammation characterized by iron deposition is observed at the edges of chronic active lesions and represents a relevant substrate of disease progression in MS. However, the influence of genetic factors on these mechanisms is not known. Leveraging the importance of iron deposition in smoldering inflammation, we assessed whether variants in genes belonging to iron-related pathways affect disease progression in MS.

**Methods:** We investigated the association between Single Nucleotide Polymorphisms (SNPs) mapping to 334 genes in iron-related pathways and the risk of disease progression, studying 2,817 MS patients from Italy (n=755) and Sweden (n=2,062), and comparing relapsing-remitting (RR-MS) with secondary progressive (SP-MS) disease course. To better understand the link of the identified variant with smoldering inflammation, we applied a multilayered approach using independent cohorts from Italy, Sweden and the United Kingdom and encompassing gene expression, PRL analysis, neurofilament levels, post-mortem spinal cord pathology and pharmacogenomics.

**Results:** We found an association between a locus in the Hypoxia-Inducible Factor 1-alpha (*HIF1A*) gene and the odds of SP-MS transition in the Italian cohort (rs11621525; SP-MS OR 0.57, 95% CI 0.44-0.72; P=3.30×10^-6^), which was replicated in the Swedish dataset (rs1951795; OR 0.79, 95% CI 0.67-0.95; P=0.0079). Additional analyses showed that patients carrying the protective allele exhibited reduced *HIF1A* expression in the immune cells, lower PRL volume, lower plasma/cerebrospinal fluid neurofilament levels, and lower inflammation and acute axonal injury in the post-mortem spinal cord. Moreover, the variant influenced the response to dimethyl fumarate, an approved MS drug with effect on mechanisms shared with *HIF1A* pathway.

**Conclusion:** A novel locus in the *HIF1A* gene, a crucial hub for iron-binding capacity, inflammation, and hypoxia response, is associated with the risk of disease progression in MS. Converging lines of evidence support the role of this locus in smoldering inflammation, prompting future studies to explore the potential of *HIF1A* as a therapeutic target in progressive MS.

## Background

Multiple sclerosis (MS) is a chronic immune-mediated degenerative disorder of the central nervous system and a leading cause of disability in young people^1^. At onset, most patients present with a relapsing-remitting course (RR-MS), typically characterized by the occurrence of relapses, followed by a variable degree of recovery^2^. Within 15-20 years from onset, the majority of people with RR-MS transitions to a secondary progressive course (SP-MS), experiencing progressive worsening of disability often independent of new relapses or new lesions at Magnetic Resonance Imaging (MRI)^2^.

Despite the high efficacy in reducing the risk of new relapses in RR-MS, the therapeutic options for progressive MS are limited by the poor understanding of the biological underpinnings of progression and the absence of *in vivo* biological markers to timely identify and track disease progression^3^.

A chronic low-grade demyelinating inflammatory process characterized by iron deposition is observed at the edges of the chronic active (or mixed active-inactive) lesions in MS at post-mortem pathology. This so-called ‘smoldering inflammation’ is thought to be a relevant mechanism for disease progression, likely acting early in the disease course^4^. Leveraging the paramagnetic properties of iron, recent MRI advances have led to the identification of the Paramagnetic Rim Lesions (PRL), an *in vivo* proxy of the chronic active lesions seen pathologically^5,6^. The PRL are correlated with chronic white matter inflammation and neuroaxonal damage, and with more pronounced disability progression^7^. For this reason, identifying and tracking the PRL using iron-sensitive MRI is under consideration as a novel endpoint in clinical trials to tackle smoldering inflammation in MS^7^.

Studying the impact of genetic variation is a fast, cost-effective, and powerful way to prioritize key molecular players in a candidate biological mechanism and pave the way to successful drug development^8^. On this basis, leveraging iron deposition as a key feature of smoldering inflammation in MS, we adopted a hypothesis-driven approach to investigate whether variants in genes belonging to iron-related pathways can influence disease progression in MS and explore the link with smoldering inflammation. To achieve this aim, we used a unique multi-layer approach that combines genetics, imaging, fluid biomarkers and quantitative neuropathological outcomes.

## Methods

### Genetic association study on iron metabolism genes

The study was conducted on a discovery Italian cohort from IRCCS San Raffaele Scientific Institute in Milan (ITA), with replication of significant associations in a Swedish nationwide cohort collected at the Karolinska Institutet in Stockholm (SWE). We compared people with RR-MS, who remained free from disease progression at over 20 years from disease onset with an Expanded Disability Status Scale (EDSS) score ≤3.5 to those with SP-MS, who had confirmed disease progression within 20 years from disease onset and an EDSS ≥4.0 at last follow-up (Table 1, Fig. 1). To provide a comprehensive set of iron-related genes, we manually screened Gene Ontology and Human Phenotype terms^9^, and the Kyoto Encyclopedia of Genes and Genome ‘*Ferroptosis*’ pathway^10^, identifying a total of 334 genes. The list of selected pathways and genes is shown in eTables 1-2 in the Supplement. Starting from whole-genome genotyping data in the discovery cohort, we extracted a total of 23,019 imputed Single Nucleotide Polymorphisms (SNPs) mapping to the selected genes, and we tested the association between these genetic variants and the course of MS, fitting logistic regression models, adjusted for biological sex and the first eight principal components, to account for population stratification. Estimation of significance level in the discovery cohort was performed applying a Bonferroni correction to the number of independent tests, considered as the number of linkage disequilibrium (LD) blocks (P<6.04×10^-5^). Statistically significant associations were then replicated in the SWE collection (Fig. 1). PLINK v1.9 software was used for quality control and the association analysis, hypothesizing a genetic additive model^11^. In the additional experiments described below, we grouped the subjects who were heterozygous or homozygous for the minor allele of the identified variant and compared them to homozygous subjects for the major allele (i.e., TT versus AT/AA), to prevent potential issues related to a low number of homozygous subjects. Detailed information is reported in the eMethods in the Supplement.

**Figure 1.**
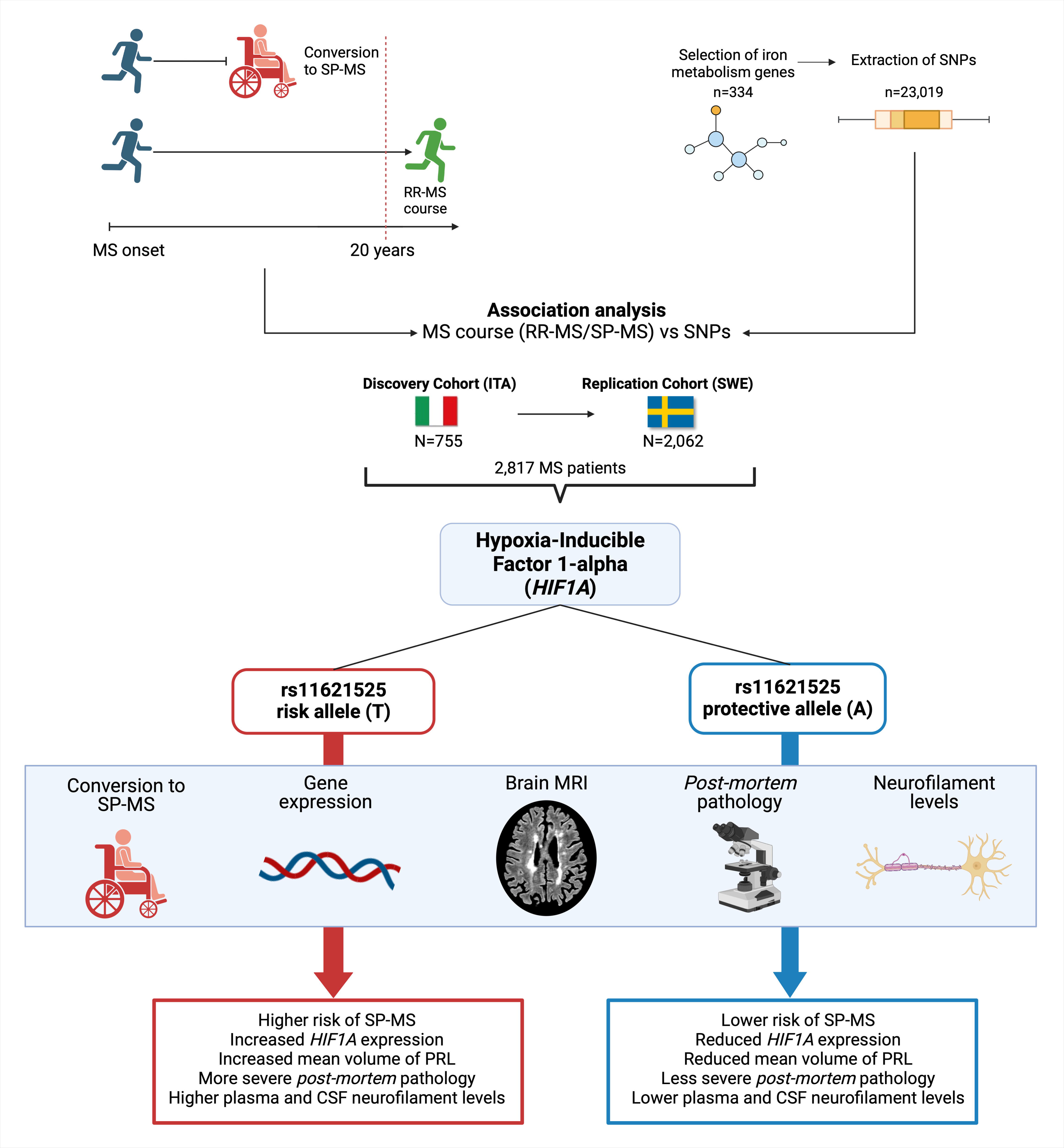
Overview of the study and main results. Patients with different clinical course in terms of disability accumulation were compared in the candidate genetic study (top left). Genes relevant for iron metabolism were manually screened across different repositories, and SNPs (n=23,019) were extracted from these genes (top right). An association analysis was then conducted on the selected genetic variants, comparing benign RR-MS and SP-MS, in a discovery cohort (ITA), with replication in a larger cohort (SWE), for a total of 2,817 MS patients. This analysis led to the identification of a variant in the *HIF1A* gene, that affects the risk of SP-MS conversion. In the bottom of the figure, we briefly summarize the effects of the protective A allele and the risk T allele, that link the SNP in HIF1A with smoldering inflammation at multiple levels (light blue panel).

**Table 1.** Clinical and demographic characteristics of the Italian discovery (ITA) and Swedish replication (SWE) cohorts involved in the genetic association study. Post-quality controls numbers are reported. Abbreviations: N = number of subjects; M = male; F = female. For course and sex, raw numbers and percentages are shown. For age at onset, the mean value (and standard deviation) of the entire cohort and of RR-MS and SP-MS patients is reported. For EDSS score at the latest follow-up, the median value (and its range) in RR-MS and SP-MS patients is reported.

|  | **ITA** | **SWE** |
| --- | --- | --- |
| **No.** | 755 | 2,062 |
| **Course** | RR-MS: 393 (52%)<br>SP-MS: 362 (48%) | RR-MS: 863 (42%)<br>SP-MS: 1199 (58%) |
| **Sex** | M: 251 (33%)<br>F: 504 (67%) | M: 539 (26%)<br>F: 1523 (74%) |
| **Age at onset** | All: 29.7 (9.68)<br>RR-MS: 26.51 (8.0)<br>SP-MS: 32.57 (10.37) | All: 32.58 (10.04)<br>RR-MS: 28.4 (7.98)<br>SP-MS: 35.6 (10.30) |
| **EDSS score at last follow-up** | RR-MS: 1.5 (0-3.5)<br>SP-MS: 6.5 (4-9.5) | RR-MS: 2 (0-3.5)<br>SP-MS: 6.5 (4-9.5) |

### Genetic influence on gene expression

To assess the regulatory effect of the identified variant on gene expression, starting from a larger cohort with whole-genome genotype data at IRCCS San Raffaele Scientific Institute, we selected an independent cohort of RR-MS patients naïve to disease-modifying treatments at sampling and with no corticosteroid exposure for at least 30 days prior to sampling. The transcriptomic profile of Peripheral Blood Mononuclear Cells (PBMC) was obtained via RNA-seq technology using the Illumina TruSeq Stranded mRNA kit. Effect of genotype on normalized gene counts was assessed via linear regression, adjusting for sex and age at sampling. Further details are described in the eMethods in the Supplement.

### PRL analysis

Brain MRI scans from MS patients with available genotype data were acquired separately in Milan and Stockholm. The PRL were identified by consensus among independent expert reviewers, as discrete fluid-attenuated inversion recovery hyperintense lesions that were either completely or partially surrounded by a paramagnetic rim of hypointense signal in unwrapped phase images (Fig. 2). After lesion identification, PRL volumes were extracted, and rank-based inverse transformed to fit a normal distribution. The impact of genotype on the lesion mean volume was first assessed independently in the two cohorts through linear regression, adjusting for disease duration, and then meta-analyzed with a fixed-effect model. See eMethods in the Supplement for additional information.

**Figure 2.**
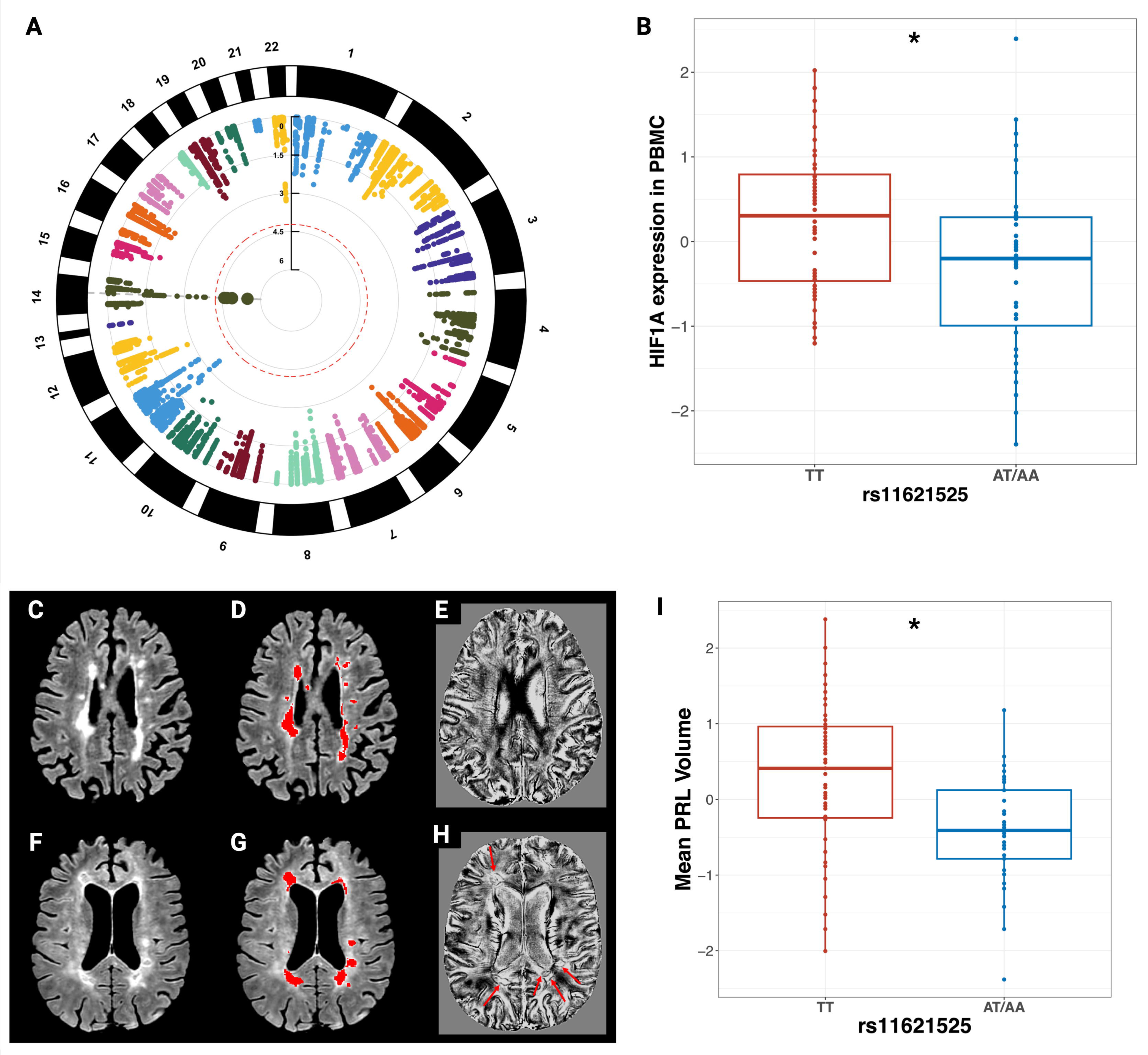
A locus in *HIF1A* is associated with the conversion to SP-MS and impacts the mean volume of the Paramagnetic Rim Lesions (PRL). (A) Circular Manhattan plot showing the results of the genetic association study. The red-dashed line marks statistical significance after correction for multiple testing (P<6.04×10^-5^). (B) The rs11621525 A allele is associated with reduced *HIF1A* expression in blood immune cells from MS patients. Distribution of the z-score of normalized *HIF1A* counts is reported for carriers of the TT genotype versus carriers of the A allele for rs11621525. (C, F) On 3D axial fluid-attenuated inversion recovery sequence, multiple T2-hyperintense white matter lesions are visible in two representative MS patients. (D, G) Brain T2-hyperintense white matter lesions (red-coded) were identified. On unwrapped phase images, no PRL were found in MS patient #1 (E), whereas several PRL (red arrows) were found in MS patient #2 (H). (I) The distribution of the z-score of normalized pooled mean PRL volumes (y-axis) is reported for individuals stratified according to the rs11621525 genotype. The mean volume of the PRL is lower in patients carrying the rs11621525 A allele. * = P<.05.

### Post-mortem pathology

Human post-mortem spinal cord material derived from a cohort of pathologically confirmed MS cases from the UK MS Tissue Bank for whom DNA sample were available. Formalin-fixed paraffin-embedded 6 µm thick adjacent sections were immunostained using primary antibodies for myelin (PLP, BioRad, #MCA839G), microglia-macrophage (CD68, Dako, #M087601-2), and acute axonal injury (BAPP, Invitrogen, #130200). Staging of white matter lesions (active, mixed active/inactive, and inactive)^12^, quantification of microglia-macrophage inflammation and neuroaxonal damage in lesional and non-lesional white matter were performed^13^. All cases were genotyped for the identified variant using a custom-made TaqMan SNP Genotyping Assay (ThermoFisher Scientific). Association between genotype and microstructural features was tested using generalized estimating equations. Additional details are reported in the eMethods in the Supplement.

### Fluid biomarkers

Plasma neurofilament light chain (NFL) levels were measured on samples collected at the Karolinska Institutet for previous studies on post-marketing treatment monitoring^14^. Data on cerebrospinal fluid (CSF) levels of NFL in patients diagnosed with MS or Clinically Isolated Syndrome (CIS) were obtained at the Karolinska Institutet from the Stockholm Prospective assessment of Multiple Sclerosis (STOP-MS) cohort, as previously described^15^. From these two datasets, we extracted subjects from the SWE cohort, involved in the above-mentioned genetic analysis, who had been sampled within 20 years from disease onset and had a RR-MS course at the time of sampling. Distance from relapses was considered to minimize a potential effect on NFL levels. When evaluating the effect of genotype on plasma NFL change after treatment start, we included from the available cohort all the RR-MS patients who had longitudinal samples at baseline and after 1 year from treatment start. NFL levels were used in simple or mixed-effect linear models as z-score, after rank-based inverse normal transformation, and were adjusted for relevant factors, as age and body mass index, as recommended^16^.

## Results

### A variant in the *HIF1A* gene affects the risk of disease progression

We tested the association between SNPs mapping to 334 genes in iron-related pathways and disease course in a total of 2,817 individuals, comparing patients who did not transition to SP-MS at more than 20 years after disease onset versus those who converted to SP-MS (Table 1). The analysis in the ITA cohort (n=755) revealed an association involving a locus on chromosome 14, that was statistically significant after correction for multiple testing (Fig. 2A, eTable 3). The identified locus is intronic to the Hypoxia-Inducible Factor 1-alfa (*HIF1A*) gene, a critical hub that is involved not only in the response to hypoxia and iron metabolism, but also in several immune mechanisms^17^. Specifically, the minor A allele of the lead variant (rs11621525) was associated with lower risk of transition to SP-MS, (odds ratio (OR) 0.57; 95% confidence interval (CI) 0.44–0.72; P=3.30×10^-6^; MAF=0.28). This association was replicated in the SWE cohort (n=2,062; lead variant: rs1951795; OR 0.79; 95% CI 0.67–0.95; P=.0079; MAF=0.16). The lead variant in the SWE cohort is in strong LD with rs11621525 (r^2^=0.98), representing a proxy for the same signal (rs11621525, P=.012) (eTable 4).

Studying the expression quantitative-trait-loci effect of the variant in PBMC obtained from 75 treatment-naïve RR-MS patients, we found that those with at least one copy of the A allele, which is protective towards disease progression, had lower *HIF1A* expression (regression coefficient (β), - 0.54; standard error (SE) 0.22; P=.019) (Fig. 2B), confirming the regulatory effect of rs11621525 on *HIF1A*, as reported in a previous study on whole-blood in ∼31,000 healthy controls^18^.

### The *HIF1A* variant influences smoldering inflammation in MS

The persistent smoldering inflammation in the PRL may lead to an outward expansion of these lesions over time^19^. To explore whether the variant in *HIF1A* may have an impact on this process, we acquired brain MRI scans from 121 patients from Italy and Sweden with available genetic data (Fig. 2C-H), identifying PRL in 72/121 patients (60%). In these patients, we found that the rs11621525 protective A allele was significantly associated with lower PRL mean volume, adjusting for disease duration and center (β -0.51; SE 0.25; P=.043; meta-analysis I^2^=0) (Fig. 2I). This finding may suggest that the protective A allele, that we have showed to reduce *HIF1A* activity through its influence on gene expression, can favor an earlier resolution of smoldering inflammation in the PRL, as observed with smaller lesion volumes.

To understand further the link with chronic inflammation, we studied the effect of the *HIF1A* variant on quantitative neuropathological outcomes, analyzing 159 spinal cord samples from 53 cases with progressive MS (eTable 5). Carriers of the protective rs11621525 A allele showed a decrease in CD68-positive microglia-macrophage inflammation in white matter lesions compared to cases harboring the TT genotype (β, -226899.84 positive pixels/mm2; SE 112006.77; P=.043) (Fig. 3A-C), a finding consistent across lesion stages (i.e., active, mixed active-inactive, and inactive). Additionally, rs11621525 A allele carriers had less acute axonal injury not only in white matter lesions (β, -17.18 BAPP-positive axons/mm^2^; SE 7.86; P=.029) (Fig. 3C), but also in the non-lesional white matter (β, -9.90 BAPP-positive axons/mm^2^; SE 4.93; P=.044) (Fig. 3D-E). A similar –but non-statistically significant– trend was observed for CD68-positive microglia-macrophage infiltration and axonal counts in non-lesional white matter (eTable 6).

**Figure 3.**
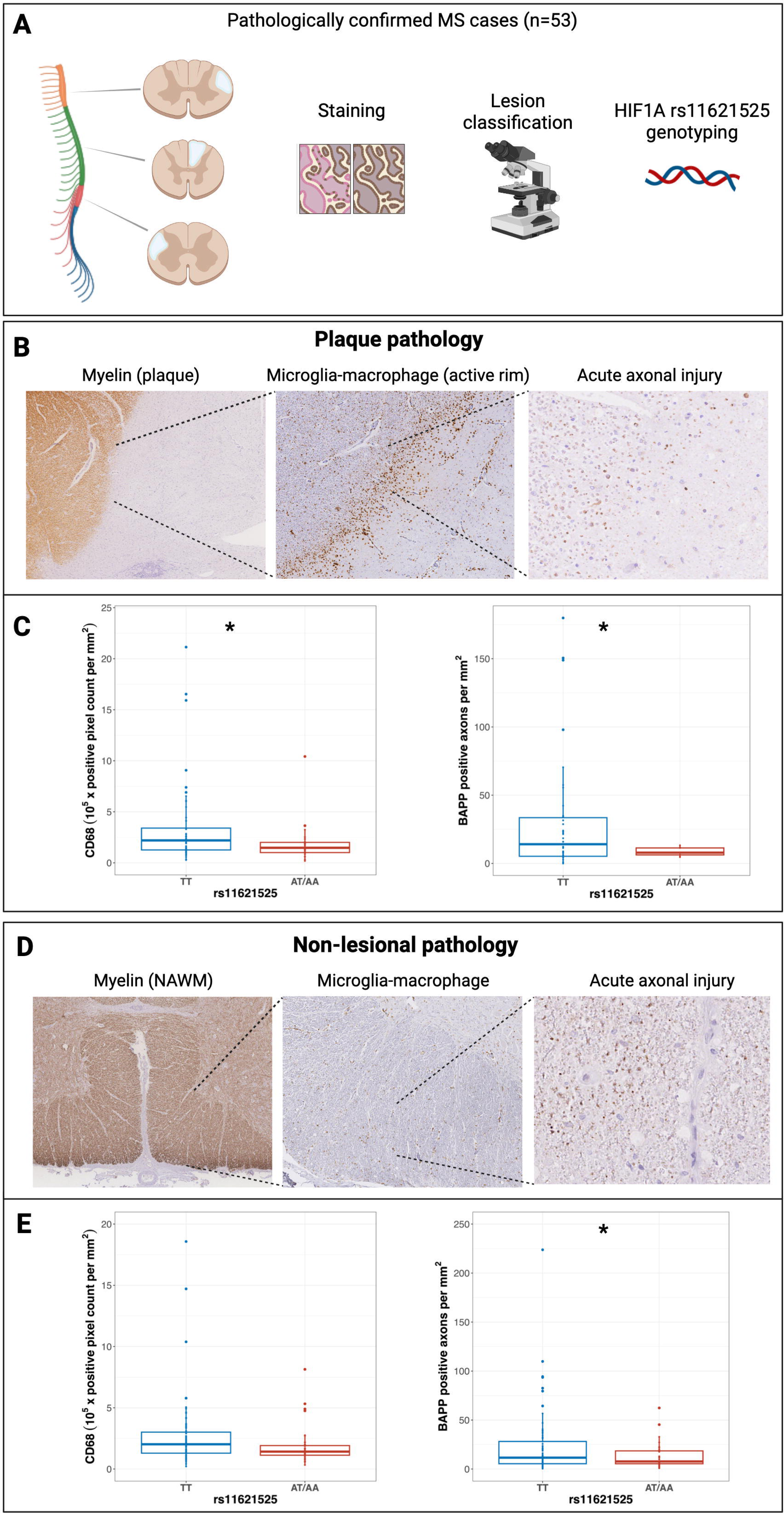
*HIF1A* genotype influences spinal cord pathology in progressive multiple sclerosis. (Panel A) Spinal cord sections from an autopsy cohort of 53 confirmed progressive MS cases were immunolabelled for myelin, microglia-macrophage, and acute axonal injury; lesion status was classified by expert reviewers. *HIF1A* rs11621525 genotype was obtained for these subjects, and its effect on microstructural pathological features was assessed. In white matter MS lesions (panels B and C), a statistically significant difference in microglia-macrophage positivity (CD68+; panel C, left boxplot) and in acute axonal injury (BAPP; panel C, right box) was found according to *HIF1A* rs11621525 genotype (TT versus AT/AA). When studying normal-appearing white matter in the spinal cord outside of MS plaques (panels D and E), the same trend was found, with patients harboring the A allele having less severe acute axonal injury (statistically significant; panel E, right boxplot) and less microglia-macrophage infiltration (non-statistically significant; panel E, left boxplot). * = P<.05; NAWM = Normal-Appearing White Matter.

### The *HIF1A* variant impacts plasma and CSF NFL levels

NFL levels, a marker of ongoing axonal injury and chronic white matter inflammation, can predict future disease progression in MS^20^. To detect whether the *HIF1A* genotype impacts early subtle inflammation and neurodegeneration mirrored by NFL levels, we studied 117 subjects from the SWE cohort that had undergone NFL measurement during the RR-MS course. Carriers of the protective allele showed lower plasma NFL levels (β, -0.54; SE 0.20; P=.0075) (Fig. 4A), with a same effect observed when measuring CSF NFL levels measured at the time of the diagnostic work-up in 71 RR-MS patients (β, -0.62; SE 0.31; P=.048) (Fig. 4B), and in a small cohort of 34 people with CIS (β, -0.74; SE 0.30; P=.019) (eFigure 1 in the Supplement).

**Figure 4.**
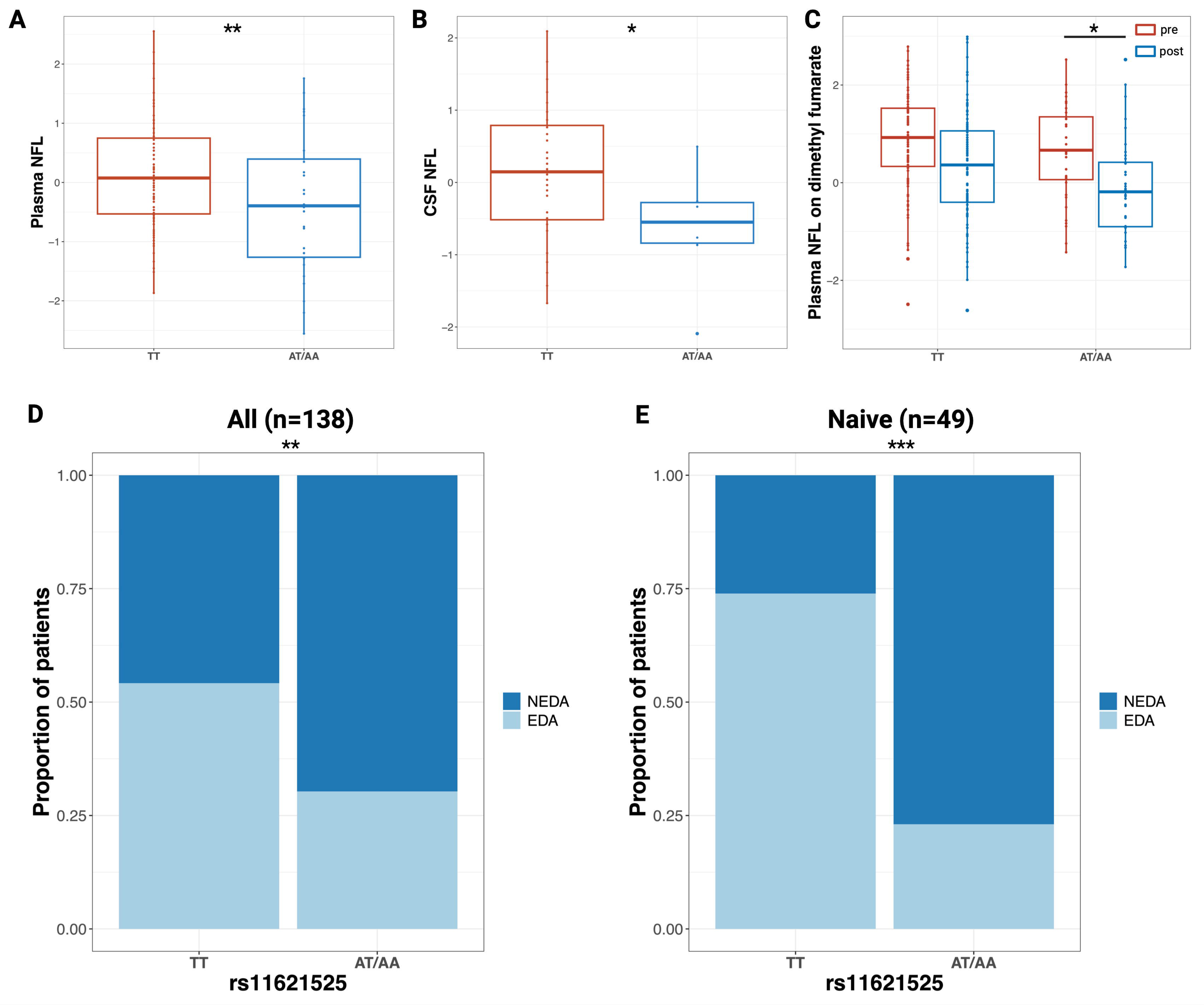
Genetic variation in *HIF1A* affects NFL levels and is associated with the response to Dimethyl Fumarate. In panels A and B, the z-scores of normalized plasma (A) and CSF (B) NFL levels are plotted on the y-axis and the genotype of rs1951795 (tagging rs11621525) is plotted on the x-axis. Panel C: the z-scores of plasma NFL levels before (“pre”; red boxplots) and after (“post”; blue boxplots) dimethyl fumarate treatment are shown according to rs1951795 genotype, with patients carrying the A allele (“AT/AA” group on the x-axis) showing a greater reduction of NFL after treatment start over time compared to the TT group (* = P-value for the effect of the SNP on NFL levels from mixed-effect linear models <.05). In panels D (all patients) and E (treatment-naïve patients), the y-axis shows the distribution of patients according to the NEDA (No Evidence of Disease Activity) and EDA (Evidence of Disease activity) status as a ratio (ranging from 0 to 1); on the x-axis the genotype for rs11621525 is shown. * = P<.05; ** = P< .01; *** = P<.001.

### Genetic variation in *HIF1A* is associated with response to dimethyl fumarate

NFL levels are known to decrease after starting a disease-modifying treatment^16^. To explore a potential link between *HIF1A* and currently approved MS treatments, we analyzed longitudinal plasma NFL measurements obtained before and after the start of dimethyl fumarate (n=151), teriflunomide (n=52), fingolimod (n=130) or natalizumab (n=178) treatment. We found that *HIF1A* A allele is associated with a more pronounced reduction of plasma NFL levels only after exposure to dimethyl fumarate (β, -0.38; SE 0.17; P=.027; Fig. 4C, eFigure 2 and eTable 7). The effect of the variant on the response to dimethyl fumarate was also confirmed at the clinical level, when evaluating 138 Italian RR-MS patients followed-up for 2 years from treatment start. Carriers of the protective A allele were more likely to achieve the No Evidence of Disease Activity status^21^ (i.e. free from relapses, disability progression and new/enlarging or gadolinium enhancing lesions at brain MRI scan), adjusting for sex and age at MS onset (OR 0.37; 95% CI 0.18–0.75; P=.0062) (Fig. 4D). This association was even stronger when restricting the analysis to the subset of patients that were treatment-naïve before dimethyl fumarate start (n=49; OR 0.083; 95% CI 0.02–0.32; P<.001) (Fig. 4E).

## Discussion

Treatment of disease progression is one of the greatest unmet needs in MS^1^. Understanding disease mechanisms is a crucial step to foster new therapeutic approaches. Recently, a multicenter effort has yielded the discovery of a variant associated with disease severity in MS^22^, highlighting the potential of genetic studies in revealing disease mechanisms. Moreover, in genetic association studies, an a priori candidate approach strongly reduces the risk of false positives due to statistical power, and significantly increases the chances of highlighting meaningful molecular targets. With these premises, in the effort of prioritizing targets for future drug development^8^ we conducted a candidate genetic association study to assess whether variants in genes belonging to iron-related pathways can influence disease progression, leveraging iron as an important feature of smoldering inflammation. To accomplish this, we took advantage of deeply characterized cohorts with long follow-up, that are highly challenging to assemble within consortium studies. To date, no study has yielded robust associations between genetic variants and the risk of transition to SP-MS, and no study has specifically investigated genes involved in processes related to smoldering inflammation and disease progression.

Our genetic analysis of two independent datasets identified a novel robust association between a variant in the *HIF1A* gene and the risk of transition to SP-MS. Specifically, carriers of the *HIF1A* protective allele, which results in decreased *HIF1A* expression in the immune cells, are more likely to have a non-progressive disease course even at long-term follow-up. The 1000 Genomes Phase 3 project reports that the protective A allele has a frequency of 31% in the general population^23^, suggesting that the contribution of the variant to the risk of progression may be relevant for a significant proportion of people with MS.

In our study, converging findings from a multi-layer set of data show the relevance of the *HIF1A* variant throughout the disease course, from CIS to end-stage. Carriers of the protective A allele had lower CSF NFL levels measured at the time of the diagnostic work-up for MS or CIS, and lower plasma NFL levels independent of future disease course. Moreover, they exhibited decreased mean volume of the PRL at brain MRI, and reduced microglia/macrophage-associated inflammation and acute axonal injury in white matter lesions in the post-mortem spinal cord, an anatomical location in which the use of iron-sensitive MRI is technically challenging. Interestingly, we found a similar effect of *HIF1A* genotype on the spinal cord matter not affected by MS lesions, in line with previous evidence demonstrating that high iron content and associated oxidative injury and neurodegeneration are prominently seen in non-lesional MS pathology^24^.

*HIF1A* has many functions, but it is mainly known as a major driver of the cell response to hypoxia. Modulation of the iron-binding capacity is an essential defense mechanism in presence of low oxygen concentration^25^, hence we can explain the role of *HIF1A* in iron-related mechanisms and its presence in the selected genes. In the context of MS, the gene is interesting not only for its functions in hypoxia and iron-binding capacity, but also as a modulator of the inflammatory response^17^. *HIF1A* is a main driver of the Th17/T regulatory cells balance, and *HIF1A*-knockout mice do not develop the Experimental Autoimmune Encephalitis, a preclinical model of MS^26^. When trying to relate this evidence to smoldering inflammation, it is known that in pathological immunological niches, as smoldering inflammation, increased energy demand may lead to a state of ‘virtual hypoxia’, a process which has already been shown in MS^27^. The virtual hypoxia can then drive the dysregulation of immune cells, mainly via transcriptional changes orchestrated by the hypoxia-inducible factors^28^. In this microenvironment, genetic factors could be relevant to limit or – conversely– maintain the chronic inflammatory process, and result in individual predisposition to develop disease progression. Our findings showing that carriers of the protective *HIF1A* allele, that results in lower *HIF1A* expression, are less prone to chronic smoldering inflammation and have significantly lower risk of disease progression, are in line with this hypothesis.

Additional evidence supporting this interpretation comes from an existing single-nuclei RNA sequencing study, that identified *HIF1A* as one of the top overexpressed genes in the chronic active edge of MS lesions^29^. In particular, in this study *HIF1A* and related genes were significantly overexpressed in a cluster of immune cells annotated as monocytes and monocytes-derived dendritic cells exposed to a hypoxic environment^29^. In addition, this observation suggests that myeloid cells could be relevant players in the mechanism involving *HIF1A*, in line with recent data showing that B lymphocyte depletion with anti-CD20 drugs was unsuccessful in limiting chronic inflammation^30^, and with the data presented herein on the effect of *HIF1A* on microglia-macrophage inflammation. Of note, HIF1A recently emerged as a main driver of the microglial dysfunction in Alzheimer’s disease and leads to Aβ plaque-associated neuropathology, providing additional evidence that the discussed mechanisms are relevant for neurodegeneration, and may be shared among different diseases^31^.

The key role of myeloid cells in smoldering inflammation in MS is also prompted by our findings that link the *HIF1A* protective allele with the response to dimethyl fumarate, in terms of lower plasma NFL levels and higher proportion of disease activity-free patients. The mechanism of action of dimethyl fumarate is not completely understood, but it is thought to regulate the hypoxic/oxidative stress response in myeloid and monocyte-derived cells, a process which is strictly linked with *HIF1A*^32^. In the same direction, a recent study reported that dimethyl fumarate is able to reduce inflammation in chronic active MS lesions^33^, a finding which requires further investigation to clarify its potential impact on disease progression.

Altogether, combining our findings and the existing evidence it seems reasonable to speculate that modifiable (e.g., lifestyle factors, drugs, comorbidities) and non-modifiable (i.e., genetic predisposition) factors with an effect on *HIF1A*-related pathways may be relevant in the development of smoldering inflammation. Future mechanistic studies are needed to better investigate this hypothesis.

In conclusion, we discovered a variant in the *HIF1A* gene that affects the risk of disease progression in MS. A unique set of clinical, radiological, fluid biomarkers and neuropathological data converge to link *HIF1A* with smoldering inflammation in MS, and therefore support the gene as a relevant target for future investigations aimed at tackling disease progression in MS.

## Data Availability

All data produced in the present are available upon reasonable request to the authors.

## Authors contribution

AG, MS, EM, SS, FC, LF, and FE contributed to the study concept or design. AG, PS, MP, MS, EM, SS, FC, LF, MC, KS, SBK, AH, FP, LA, JH, TO, KA, IK, and FE contributed to collection or elaboration of the genetic data. PP, RO, FP, ES, TG, MR, and MF were involved in the acquisition or elaboration of MRI data. MP and GCD contributed to analysis of pathology data. AG, AM, TM, FP, LA, JH, TO, and IK were involved in collection or elaboration of the neurofilament data. AG and MP performed the statistical analyses. AG, PS, PP, MP, KM, AM, TM, KS, RO, ES, IK, FE had access to raw data. AG, PS, PP, MP, AM, and TM verified the data. MN, AM, FP, TG, LF, MJ, GCD, MR, MF, IK, and FE provided supervision. All authors contributed to critical review and had final responsibility for the decision to submit for publication.

## Funding and support

This study was supported by ERA PerMed-JTC2018 (co-funded by the European Commission) and the European Union in the “Horizon 2020 – Framework Programme for Research and Innovation (2014-2020)”.

## Role of the Funders

The funding sources had no role in the design and conduct of the study; collection, management, analysis, and interpretation of the data; preparation, review, or approval of the manuscript; and decision to submit the manuscript for publication.

## Declaration of interest

AG, PS, MP, MS, EM, SS, KM, FC, MN, MC, TM, KS, RO, AH, ES, SBK, and KA: nothing to disclose. PP reports compensation for speaking activity from Bristol Myers Squibb, Sanofi Genzyme, Novartis, Roche, Merck. LF received intellectual property interests from a discovery or technology relating to health care. AM reports compensation for consulting services from Biogen and for speaking activity from Biogen, research support from Karolinska Institutet. FP reports compensation for participation in advisory boards from Parexel/Chugai, Swedish Medical Products Agency, research support from UCB, Merck. LA reports research support from AFA insurance, Swedish Research Council, Swedish Brain Foundation, Region Stockholm. JH reports compensation for participation in advisory boards from Biogen, Celgene, Merck, Novartis, Sandoz, Sanofi, research support from Biogen, Celgene, Merck, Novartis, Sanofi, and Roche. TO reports compensation for participation in advisory boards from Biogen, Novartis, Merck, Sanofi, research support from Biogen, Novartis, Sanofi, Merck. TG reports research support from Karolinska Institutet, Region Stockholm, Merck, Swedish Society for Medical Research. MJ reports research support from Swedish Research Council, EU Horizon, European Research Council (ERC-CoG). GCD reports compensation for consulting services from Neurology Academy, for speaking activity from Merck, Roche, research support from NIHR, BRC (Oxford), National Health and Medical Research (Australia), UK MS Society, Oxford-Quinnipiac Partnership, US Department of Defense, Wellcome ISSF, Bristol Myers Squibb, University of Oxford (John Fell Fund), for serving as an editorial board member for MS Journal. MR received consulting fees from Biogen, Bristol-Myers Squibb, Eli Lilly, Janssen, and Roche; speaker honoraria from AstraZeneca, Biogen, Bristol-Myers Squibb, Bromatech, Celgene, Genzyme, Horizon Therapeutics Italy, Merck-Serono, Novartis, Roche, Sanofi, and Teva Pharmaceuticals; and research support from the Multiple Sclerosis Society of Canada, the Italian Ministry of Health, and Fondazione Italiana Sclerosi Multipla. MF reports compensation for consulting services from Alexion, Almirall, Biogen, Merck, Novartis, Roche, and Sanofi; speaking activities from Bayer, Biogen, Celgene, Chiesi Italia, Eli Lilly, Genzyme, Janssen, Merck-Serono, Neopharmed Gentili, Novartis, Novo Nordisk, Roche, Sanofi, Takeda, and Teva; participation in advisory boards for Alexion, Biogen, Bristol-Myers Squibb, Merck, Novartis, Roche, Sanofi, Sanofi-Aventis, Sanofi-Genzyme, and Takeda; scientific direction of educational events for Biogen, Merck, Roche, Celgene, Bristol-Myers Squibb, Eli Lilly, Novartis, and Sanofi-Genzyme; and research support from Biogen Idec, Merck-Serono, Novartis, Roche, Italian Ministry of Health, and Fondazione Italiana Sclerosi Multipla. IK reports compensation for speaking activity from Merck, research support from Swedish Research Council, National MS society, EU Horizon 2020. FE reports compensation for consulting services from Merck, Novartis, for participation in advisory boards from Novartis, Merck, research support from Italian MS Society, Italian Ministry of Health, ERA Net, European Commission. FE has received intellectual property interests from a discovery or technology relating to health care.

## Notes

### Competing Interest Statement

AG, PS, MP, MS, EM, SS, KM, FC, MN, MC, TM, KS, RO, ES, AH, SBK, and KA: nothing to disclose. PP reports compensation for speaking activity from Bristol Myers Squibb, Sanofi Genzyme, Novartis, Roche, Merck. LF received intellectual property interests from a discovery or technology relating to health care. AM reports compensation for consulting services from Biogen and for speaking activity from Biogen, research support from Karolinska Institutet. FP reports compensation for participation in advisory boards from Parexel/Chugai, Swedish Medical Products Agency, research support from UCB, Merck. LA reports research support from AFA insurance, Swedish Research Council, Swedish Brain Foundation, Region Stockholm. JH reports compensation for participation in advisory boards from Biogen, Celgene, Merck, Novartis, Sandoz, Sanofi, research support from Biogen, Celgene, Merck, Novartis, Sanofi, and Roche. TO reports compensation for participation in advisory boards from Biogen, Novartis, Merck, Sanofi, research support from Biogen, Novartis, Sanofi, Merck. TG reports research support from Karolinska Institutet, Region Stockholm, Merck, Swedish Society for Medical Research. MJ reports research support from Swedish Research Council, EU Horizon, European Research Council (ERC-CoG). GCD reports compensation for consulting services from Neurology Academy, for speaking activity from Merck, Roche, research support from NIHR, BRC (Oxford), National Health and Medical Research (Australia), UK MS Society, Oxford-Quinnipiac Partnership, US Department of Defense, Wellcome ISSF, Bristol Myers Squibb, University of Oxford (John Fell Fund), for serving as an editorial board member for MS Journal. MR received consulting fees from Biogen, Bristol-Myers Squibb, Eli Lilly, Janssen, and Roche; speaker honoraria from AstraZeneca, Biogen, Bristol-Myers Squibb, Bromatech, Celgene, Genzyme, Horizon Therapeutics Italy, Merck-Serono, Novartis, Roche, Sanofi, and Teva Pharmaceuticals; and research support from the Multiple Sclerosis Society of Canada, the Italian Ministry of Health, and Fondazione Italiana Sclerosi Multipla. MF reports compensation for consulting services from Alexion, Almirall, Biogen, Merck, Novartis, Roche, and Sanofi; speaking activities from Bayer, Biogen, Celgene, Chiesi Italia, Eli Lilly, Genzyme, Janssen, Merck-Serono, Neopharmed Gentili, Novartis, Novo Nordisk, Roche, Sanofi, Takeda, and Teva; participation in advisory boards for Alexion, Biogen, Bristol-Myers Squibb, Merck, Novartis, Roche, Sanofi, Sanofi-Aventis, Sanofi-Genzyme, and Takeda; scientific direction of educational events for Biogen, Merck, Roche, Celgene, Bristol-Myers Squibb, Eli Lilly, Novartis, and Sanofi-Genzyme; and research support from Biogen Idec, Merck-Serono, Novartis, Roche, Italian Ministry of Health, and Fondazione Italiana Sclerosi Multipla. IK reports compensation for speaking activity from Merck, research support from Swedish Research Council, National MS society, EU Horizon 2020. FE reports compensation for consulting services from Merck, Novartis, for participation in advisory boards from Novartis, Merck, research support from Italian MS Society, Italian Ministry of Health, ERA Net, European Commission. FE has received intellectual property interests from a discovery or technology relating to health care.

### Author Declarations

Ethics committee of IRCCS San Raffaele Scientific Institute, Karolinska Institutet and Oxford Neuroinflammation Center gave ethical approval for this work. Approval numbers are as follows. IRCCS San Raffaele Scientific Institute: 1013/DG, 274/DG, 397/ER/mm, 851/DG, 20/int/2019. Karolinska Institutet: 02-548, 2006/845-31/1, 2009/2107-31/2. 2011/641-31/4, 02-548, 2015/2235-32/4, 2015/2236-32, 2015/2260-32/2, 2016/1167-32, 2016/1168-32, 2016/1169-32, 2017/1347-32, 2017/1348-32, 2017/1349-32, 2017/1350-32, 2017/1426-32. Oxford Neuroinflammation Center: REC-08/MRE09/31+5.

## REFERENCES

1. Bebo B, Cintina I, LaRocca N, et al. The Economic Burden of Multiple Sclerosis in the United States: Estimate of Direct and Indirect Costs. Neurology. 2022;98(18):e1810–e1817. doi:10.1212/WNL.0000000000200150

2. Confavreux C, Vukusic S, Moreau T, Adeleine P. Relapses and Progression of Disability in Multiple Sclerosis. N Engl J Med. 2000;343(20):1430–1438. doi:10.1056/NEJM200011163432001

3. Yong HYF, Yong VW. Mechanism-based criteria to improve therapeutic outcomes in progressive multiple sclerosis. Nat Rev Neurol. 2022;18(1):40–55. doi:10.1038/s41582-021-00581-x

4. Giovannoni G, Popescu V, Wuerfel J, et al. Smouldering multiple sclerosis: the ‘real MS.’ Ther Adv Neurol Disord. 2022;15:175628642110667. doi:10.1177/17562864211066751

5. Absinta M, Sati P, Schindler M, et al. Persistent 7-tesla phase rim predicts poor outcome in new multiple sclerosis patient lesions. J Clin Invest. 2016;126(7):2597–2609. doi:10.1172/JCI86198

6. Absinta M, Sati P, Fechner A, Schindler MK, Nair G, Reich DS. Identification of Chronic Active Multiple Sclerosis Lesions on 3T MRI. Am J Neuroradiol. 2018;39(7):1233–1238. doi:10.3174/ajnr.A5660

7. Preziosa P, Filippi M, Rocca MA. Chronic active lesions: a new MRI biomarker to monitor treatment effect in multiple sclerosis? Expert Rev Neurother. 2021;21(8):837–841. doi:10.1080/14737175.2021.1953983

8. Plenge RM, Scolnick EM, Altshuler D. Validating therapeutic targets through human genetics. Nat Rev Drug Discov. 2013;12(8):581–594. doi:10.1038/nrd4051

9. Ashburner M, Ball CA, Blake JA, et al. Gene Ontology: tool for the unification of biology. Nat Genet. 2000;25(1):25–29. doi:10.1038/75556

10. Kanehisa M. KEGG: Kyoto Encyclopedia of Genes and Genomes. Nucleic Acids Res. 2000;28(1):27–30. doi:10.1093/nar/28.1.27

11. Chang CC, Chow CC, Tellier LC, Vattikuti S, Purcell SM, Lee JJ. Second-generation PLINK: rising to the challenge of larger and richer datasets. GigaScience. 2015;4(1):7. doi:10.1186/s13742-015-0047-8

12. Kuhlmann T, Ludwin S, Prat A, Antel J, Brück W, Lassmann H. An updated histological classification system for multiple sclerosis lesions. Acta Neuropathol (Berl). 2017;133(1):13–24. doi:10.1007/s00401-016-1653-y

13. DeLuca GC. The contribution of demyelination to axonal loss in multiple sclerosis. Brain. 2006;129(6):1507–1516. doi:10.1093/brain/awl074

14. Hillert J, Stawiarz L. The Swedish MS registry – clinical support tool and scientific resource. Acta Neurol Scand. 2015;132(S199):11–19. doi:10.1111/ane.12425

15. Huang J, Khademi M, Fugger L, et al. Inflammation-related plasma and CSF biomarkers for multiple sclerosis. Proc Natl Acad Sci. 2020;117(23):12952–12960. doi:10.1073/pnas.1912839117

16. Benkert P, Meier S, Schaedelin S, et al. Serum neurofilament light chain for individual prognostication of disease activity in people with multiple sclerosis: a retrospective modelling and validation study. Lancet Neurol. 2022;21(3):246–257. doi:10.1016/S1474-4422(22)00009-6

17. Tang YY, Wang DC, Wang YQ, Huang AF, Xu WD. Emerging role of hypoxia-inducible factor-1α in inflammatory autoimmune diseases: A comprehensive review. Front Immunol. 2023;13:1073971. doi:10.3389/fimmu.2022.1073971

18. Võsa U, Claringbould A, Westra HJ, et al. Large-scale cis- and trans-eQTL analyses identify thousands of genetic loci and polygenic scores that regulate blood gene expression. Nat Genet. 2021;53(9):1300–1310. doi:10.1038/s41588-021-00913-z

19. Dal-Bianco A, Grabner G, Kronnerwetter C, et al. Long-term evolution of multiple sclerosis iron rim lesions in 7 T MRI. Brain. 2021;144(3):833–847. doi:10.1093/brain/awaa436

20. Uphaus T, Steffen F, Muthuraman M, et al. NfL predicts relapse-free progression in a longitudinal multiple sclerosis cohort study. eBioMedicine. 2021;72:103590. doi:10.1016/j.ebiom.2021.103590

21. Havrdova E, Galetta S, Stefoski D, Comi G. Freedom from disease activity in multiple sclerosis. Neurology. 2010;74(Issue 17, Supplement 3):S3–S7. doi:10.1212/WNL.0b013e3181dbb51c

22. International Multiple Sclerosis Genetics Consortium, Harroud A, Stridh P, et al. Locus for severity implicates CNS resilience in progression of multiple sclerosis. Nature. 2023;619(7969):323–331. doi:10.1038/s41586-023-06250-x

23. The 1000 Genomes Project Consortium, Corresponding authors, Auton A, et al. A global reference for human genetic variation. Nature. 2015;526(7571):68–74. doi:10.1038/nature15393

24. Haider L, Simeonidou C, Steinberger G, et al. Multiple sclerosis deep grey matter: the relation between demyelination, neurodegeneration, inflammation and iron. J Neurol Neurosurg Psychiatry. 2014;85(12):1386–1395. doi:10.1136/jnnp-2014-307712

25. Peyssonnaux C, Zinkernagel AS, Schuepbach RA, et al. Regulation of iron homeostasis by the hypoxia-inducible transcription factors (HIFs). J Clin Invest. 2007;117(7):1926–1932. doi:10.1172/JCI31370

26. Dang EV, Barbi J, Yang HY, et al. Control of TH17/Treg Balance by Hypoxia-Inducible Factor 1. Cell. 2011;146(5):772–784. doi:10.1016/j.cell.2011.07.033

27. Trapp BD, Stys PK. Virtual hypoxia and chronic necrosis of demyelinated axons in multiple sclerosis. Lancet Neurol. 2009;8(3):280–291. doi:10.1016/S1474-4422(09)70043-2

28. Taylor CT, Colgan SP. Regulation of immunity and inflammation by hypoxia in immunological niches. Nat Rev Immunol. 2017;17(12):774–785. doi:10.1038/nri.2017.103

29. Absinta M, Maric D, Gharagozloo M, et al. A lymphocyte–microglia–astrocyte axis in chronic active multiple sclerosis. Nature. 2021;597(7878):709–714. doi:10.1038/s41586-021-03892-7

30. Maggi P, Bulcke CV, Pedrini E, et al. B cell depletion therapy does not resolve chronic active multiple sclerosis lesions. eBioMedicine. 2023;94:104701. doi:10.1016/j.ebiom.2023.104701

31. March-Diaz R, Lara-Ureña N, Romero-Molina C, et al. Hypoxia compromises the mitochondrial metabolism of Alzheimer’s disease microglia via HIF1. Nat Aging. 2021;1(4):385–399. doi:10.1038/s43587-021-00054-2

32. Carlström KE, Ewing E, Granqvist M, et al. Therapeutic efficacy of dimethyl fumarate in relapsing-remitting multiple sclerosis associates with ROS pathway in monocytes. Nat Commun. 2019;10(1):3081. doi:10.1038/s41467-019-11139-3

33. Zinger N, Ponath G, Sweeney E, et al. Dimethyl Fumarate Reduces Inflammation in Chronic Active Multiple Sclerosis Lesions. Neurol - Neuroimmunol Neuroinflammation. 2022;9(2):e1138. doi:10.1212/NXI.0000000000001138

