## Supplement for "Genetic variation in *HIF1A* is associated with smoldering inflammation and disease progression in Multiple Sclerosis"

**Giordano A, et al.**

### Supplement

### SUPPLEMENTARY METHODS

#### Genetic association study

##### *Cohorts for the genetic study*

For the genetic association study, we compared RR-MS patients, who did not experience disease progression for the first 20 years from disease onset and with an Expanded Disability Status Scale (EDSS) score ≤ 3.5 at last follow-up, with SP-MS patients, who had confirmed disease progression within 20 years from disease onset and EDSS ≥ 4.0 at last follow-up. Starting from a large cohort of MS patients with available whole-genome genotype data at the Laboratory of Human Genetics of Neurological Disorders at the San Raffaele Scientific Institute (ITA), we identified 772 patients with definite diagnosis of MS according to 2017 revised criteria[1], who fulfilled the above-mentioned inclusion criteria for RR-MS (n=406) or SP-MS (n=366), and underwent quality control for the genetic study. To replicate significant associations emerging from the ITA cohort study, we took advantage of a large Swedish nationwide cohort (SWE) gathered at the Karolinska Institutet (Stockholm, Sweden). The subjects of the SWE cohort were part of the National Swedish MS Registry. Clinical information and blood samples for genotyping were gathered and managed at the Karolinska Institutet. In the SWE cohort, a total of 2,062 patients with RR-MS (n=863) and SP-MS (n=1,199) were included, who fulfilled the same criteria used for the discovery cohort.

##### *Genotype imputation and quality control in the ITA cohort*

As mentioned, a large cohort of MS patients with available whole-genome genotype information was available at the Human Genetics of Neurological Disorders Unit at San Raffaele Scientific Institute. Imputation to reference genome from Haplotype Reference Consortium[2] r1.1 had been conducted on the entire cohort, using Eagle2 for phasing and Minimac4 for missing genotype imputation. Since individuals had been genotyped on four different platforms (Illumina OmniExpress, Illumina Omni 2.5, Illumina Human Quad, Illumina Global Screening Array), imputation was carried out separately on each platform with later merging on bona-fide imputed variants. Given the high level of overlap between OmniExpress and Omni 2.5, samples genotyped on these two platforms were jointly quality-controlled and imputed, retaining only markers with good imputation quality (Rsq>0.6). Variants' rsIDs were assigned from dbSNP v151 GRCh37p13 (http://www.ncbi.nlm.nih.gov/SNP/).

Prior to imputation, a set of homogeneous quality control (QC) metrics at sample and SNP level were conducted for each of the cohorts separately. At sample level, we excluded subjects for which a mismatch with declared sex was observed, those with call-rate < 90% and outliers exceeding the mean level of heterozygosity by >3 standard deviations. At variant level, we discarded rare SNPs with Minor Allele Frequency (MAF) <1%, SNPs with a call-rate<90% and those departing from Hardy-Weinberg Equilibrium (HWE) at $p < 1 \times 10^{-6}$.

From the entire available cohort, patients were screened to match the inclusion criteria for RR-MS and SP-MS and were included in the genetic association study here described. We initially identified 772 patients fulfilling the inclusion criteria, out of which we excluded 13 subjects who had not passed the imputation QC.

Additional study-specific QC was conducted on the remaining 759 subjects and 9,388,047 imputed genetic variants. First, we applied a filter for missingness ≥ 0.02 (no subject was excluded). To discover individuals with potential genetic relationship, we performed pairwise identity by descent (IBD) estimate, identifying 6 potentially related subjects (PI_HAT ≥ 0.250). For each pair, we randomly selected and removed one subject (n=3). To identify population outliers (more than ±6 standard deviations from population mean), we run a Principal Component Analysis (PCA) and excluded one subject. The final number of subjects included in the study was 755. For both IBD estimation and PCA, the regions with extended linkage disequilibrium (LD) were removed and the dataset was pruned (no pair of SNPs with $r^2 > 0.1$ in a window of 1000 kb).

Subsequently, we filtered out all the SNPs with MAF ≤ 0.05, genotyping call rate < 99% and HWE ($P<1\times10^{-6}$), for a total of 4,272,520 genetic variants in 755 subjects that were included in the study.

##### *Selection of SNPs in iron metabolism genes*

To provide a comprehensive list of genes involved in iron metabolism, we manually screened Gene Ontology and Human Phenotype terms related to iron homeostasis[3]. This list of genes was merged with the Kyoto Encyclopedia of Genes and Genome '*Ferroptosis*' pathway[4]. In total, we identified 336 unique genes relevant to iron metabolism. GRCh37/hg19 genes coordinates were extracted using UCSC Table Browser[5]. In total, 334 out of 336 genes were mapped, and the longest isoform was selected, adding a 2 kilobase (kb) flanking region both upstream and downstream of the gene to account for regulatory variation. Complete lists of the pathways and mapped genes are reported in the eTables 1 and 2. A total of 23,019 high coverage imputed SNPs mapped to the selected iron metabolism genes and underwent association analysis.

##### *Genetic association analysis*

We tested the association between the 23,019 genetic variants mapping to iron metabolism genes and the course of MS fitting logistic regression models, adjusted for biological sex and the first eight principal components to account for population stratification. An additive model was used to investigate the effect of genotype on disease course. In the additional experiments, we grouped the subjects who were heterozygous or homozygous for the minor allele of the identified variant and compared them to homozygous subjects for the major allele (i.e., TT versus AT/AA), to prevent potential issues related to a low number of homozygous subjects. PLINK v1.9 software was used for quality control and the association analysis[6].

***Estimation of significance level for genetic association analysis***

To calculate a threshold for statistical significance that was able to provide robust results despite multiple testing, we estimated the haplotype blocks in the whole-genome imputed ITA cohort data using PLINK v1.9[6,7]. We identified a total of 828 blocks in the 334 iron metabolism genes. On this number, we applied a Bonferroni correction to minimize the risk of type I error. Following this approach, the threshold for statistical significance was established at $P<6.04x\text{-}10^{-5}$ (0.05/828).

***Genotype and quality control in the SWE cohort***

Replication of the statistically significant association found in the ITA cohort was conducted in the SWE cohort. Genotyping in the SWE cohort had been performed for previous studies using the Illumina Human OmniExpress platform, carried out by deCODE Genetics (Reykjavik, Iceland). Quality control had been applied as follows. The cohort was initially aligned to the forward strand of the GRC37/hg19 reference genome, according to strand details from the Illumina array manifest files. Any samples presenting with a genotyping yield less than 95% or displaying an inconsistency between stated sex and genetic sex were excluded. This determination was based on a LD pruned set of high-quality chromosome X variants with less than 2% missingness and a MAF higher than 5%. The screening process for variants included conditions such as genotype missingness < 2%, deviation from HWE with $P<1x10^{-6}$ for non-MS individuals, MAF >1%, and a differential missingness between MS and non-MS individuals with $P>1x10^{-4}$. For palindromic variants, alternative allele frequencies were set between 0.4 and 0.6. Individuals demonstrating an absolute inbreeding coefficient > 0.05 or relatedness at the third degree or closer were not included. Before imputation, PCs were calculated, and the samples were projected onto the PC space of 1000 Genomes phase 3 samples. Any samples that deviated more than 6 standard deviations from the mean of the European superpopulation were disregarded. Genotyped data management was performed using PLINK v1.9. During imputation, phasing of 20 Mb blocks, including 5 Mb overlapping flank regions, was conducted using Eagle2 (version v2.4.1), with the number of conditioning haplotypes set to 20,000 (default 10,000). The Haplotype Reference Consortium (HRC; version 1.1) imputation reference panel was employed in the application of Minimac4 for the imputation process. Quality control was conducted by reviewing chromosome continuity, evaluating imputation quality according to chromosome position, and comparing allele frequency with the reference panel. For variants with MAF $\geq$0.01, the median imputation quality ($R^2$) ranged from 0.965 to 0.985, and the median imputation accuracy for genotyped variants (EmpR) spanned from 0.977 to 0.997. Imputed variants with MAF < 0.01 or R2 <0.3 were not included, which resulted in a total of 7,743,331 available autosomal variants.

***Replication in the SWE cohort***

From whole-genome imputed data, we extracted the subjects and SNPs relevant to our analysis. To replicate significant association found in the ITA cohort, the threshold for statistical significance was set at $p < 0.05$, given that only one signal had passed the threshold for statistical significance after multiple testing correction. As lead variant of the signal in the SWE cohort we considered the SNP having the lowest p-value among those in almost perfect linkage disequilibrium ($r^2 \geq 0.95$) with rs11621525 (eTable 4). The association analysis was conducted using logistic regression, adjusting for biological sex and the first eight principal components, as reported for the ITA cohort.

**Effect of genotype on gene expression**

***Inclusion criteria, genotyping and transcriptomic profiling***

For the study on the effect of the rs11621525 variant on *HIF1A* gene expression, inclusion criteria were: a) diagnosis of RR-MS; b) fully naive from disease-modifying treatment (DMT) at the time of sampling; c) no treatment with corticosteroid drugs in the 30 days before sampling; d) availability of rs11621525 genotype.

Genotype information for these patients had been acquired using Illumina OmniExpress beadchips, and imputation had been carried out on a larger cohort available at San Raffaele Scientific Institute, as described above.

For the RNA-sequencing experiment, after blood sampling, Peripheral Blood Mononuclear Cells (PBMC) were isolated through density gradient using Lymphoprep (StemCell Technologies) and RNA was extracted using Quick-DNA/RNA Miniprep Plus (Zymo). The transcriptomic profile was obtained via RNA-seq technology using the Illumina TruSeq Stranded mRNA kit. RNA libraries were sequenced on a HiSeq3000 sequencer (Illumina), reaching >25 million reads/samples.

***Bioinformatic pipeline and analysis***

RNA-seq reads were aligned to hg19 reference genome, removing poor-quality bases and adapter sequences[8,9]. Quantification of gene expression levels and gene-level summarization was performed using featureCounts[10], according to GENCODE v19 annotation gene model. Quality control of raw and aligned reads was performed by MultiQC tool[11]. To discard features deemed as not expressed, we only retained those with > 5 counts in at least 25% of the whole cohort.

The counts of the *HIF1A* gene were extracted, and the effect of genotype on gene expression was calculated via linear regression applying a dominant model for genotype, using normalized gene counts (z-score) as outcome and genotype as predictor, adjusting for sex and age at sampling.

**Effect of genotype on PRL mean volume**

***MRI acquisition***

MRI acquisition was performed separately by the Neuroimaging Research Unit at IRCCS San Raffaele Scientific Institute and by the Department of Neuroradiology at Karolinska University Hospital for patients with available genotype information (see above for detail on genetic data).

For the Italian cohort, a 3.0 Tesla Philips Ingenia CX scanner (Philips Medical Systems) was used. The following sequences were acquired: (a) sagittal three dimensional (3D) fluid attenuation inversion recovery (FLAIR), field of view (FOV)=256×256 mm, voxel size=1×1×1 mm, 192 slices, matrix=256×256, repetition time (TR)=4800 ms, echo time (TE)=270 ms, inversion time (TI)=1650 ms, echo train length (ETL)=167, acquisition time (TA)=6.15 min; (b) sagittal 3D T1-weighted turbo field echo, FOV=256×256, voxel size=1×1×1 mm, 204 slices, matrix=256×256, TR=7 ms, TE=3.2 ms, TI=1000 ms, flip angle=8°, TA=8.53 min; (c) 3D T2-weighted scan, FOV=256×256 mm, pixel size=1×1 mm, 192 axial slices with 1 mm slice thickness, TR 2500 ms, TE 330 ms, ETL=117, TA= 3 min; (c) 3D susceptibility-weighted image (SWI), FOV=230x230, pixel size=0.60×0.60 mm, 135 slices, 2 mm-thick, matrix=384x382, TR=39 ms, TEs=5.5:6:35.5 ms, flip angle=17°, TA=6 min; both magnitude and phase images for each echo were saved.

At the Karolinska University Hospital, brain MRI were acquired using a 3.0 Tesla Siemens MAGNETOM Prisma$^{Fit}$ scanner. The following sequences were acquired: (a) sagittal 3D FLAIR, FOV=256×228 mm, voxel size=1×1×1 mm, 160 slices, matrix=256×228, TR=5000 ms, echo time TE=386 ms, TI=1600 ms, TA=4.52 min; (b) sagittal 3D T1-weighted Magnetization-Prepared Rapid Acquisition Gradient Echo (MPRAGE), FOV=256×256, voxel size=1×1×1 mm, 176 slices, matrix=256×256, TR=1900 ms, TE=2.5 ms, TI=900 ms, flip angle=9°, TA=4.26 min; (c) sagittal 3D T2-weighted (SPACE) turbo spine-echo, FOV=256×256 mm, pixel size=1×1×1 mm, 176 slices, TR 3200 ms, TE 410 ms, TA= 3.49 min; (d) A 3D SWI sequence was acquired with the following protocol: FOV=220x200, pixel size=0.90×0.90 mm, 72 slices, 2 mm-thick, matrix=244x222, TR=28 ms, TEs=20 ms, flip angle=15°, TA=4.37 min; both magnitude and phase images for each echo were saved.

***Identification of the PRL and volume extraction***

A fully automated approach using the 3D FLAIR and 3D T1weighted as input images[12] was used by Neuroimaging Research Unit at San Raffaele Scientific Institute to identify brain T2-hyperintense WM lesions. T2-hyperintense WM lesion volume was obtained for each patient from their lesion masks, after a careful visual check of the results provided by the automatic segmentation by a team of expert neurologists. The MRI team at the Department of Neuroradiology at Karolinska University Hospital similarly identified brain T2-hyperintense WM lesions using a semi-automated approach, beginning with automatic segmentation using lesion segmentation toolbox lesion prediction algorithm[13] followed by manual edits first by an experienced rater and a trained radiology resident to be finally approved together with a neuroradiologist for consensus agreement.

The PRL were defined as discrete FLAIR-hyperintense lesions either completely or partially surrounded by a paramagnetic rim of hypointense signal in unwrapped phase images.

For each patient, the number and volume of total T2-hyperintense WM lesions as well as of T2-hyperintense WM lesions with or without the hypointense paramagnetic rim were automatically estimated. Firstly, from the global lesion mask, different intensity values were manually given according to the different type of lesions (1=non-paramagnetic rim lesion, 2=paramagnetic rim lesion) creating a new label mask. Then, an automatic pipeline estimated the number and the dimension of the 3D connected objects (lesions) found within label masks, separately for each type of lesion (intensity value). Total number of T2-hyperintense WM lesions was obtained from the sum of T2-hyperintense WM lesions with or without the hypointense paramagnetic rim. Volumes were finally obtained by correcting the dimension for the voxel size and converted in milliliters (ml).

***Statistical analysis***

Given the non-normal distribution of the volumetric information (Shapiro-test $p<0.001$), the mean volumes of the PRL were rank-based inverse transformed to be suitable for testing using a linear model. The impact of genotype on the lesion mean volume was first assessed independently in the two cohorts through linear regression, adjusting for disease duration, and then meta-analyzed using a fixed-effect model.

**Spinal cord pathology**

***HIF1A genotyping, staining and lesions classification***

Cervical, thoracic, and lumbar spinal cord sections from a human autopsy cohort of pathologically confirmed MS cases (n=53) was obtained from the UK Multiple Sclerosis Tissue Bank in compliance with Human Tissue Act guidelines (REC 08/MRE09/31+5).

DNA extracted from freshly frozen cerebellar tissue from MS cases was used for *HIF1A* rs11621525 genotyping using a custom-made TaqMan SNP Genotyping Assay manufactured by ThermoFisher Scientific.

The study was conducted on 108 samples from 36 cases with TT genotype and on 51 samples from 17 cases who carried the rs11621525 A allele. Cases were matched for demographic variables, including sex, age at death, disease duration, *HLA-DRB1*15:01* status, brain weight and post-mortem interval, as shown in eTable 5.
Formalin-fixed paraffin-embedded 6 μm thick adjacent sections were immunostained using primary antibodies for myelin (PLP, BioRad, #MCA839G), microglia-macrophage (CD68, Dako, #M087601-2), and acute axonal injury (BAPP, Invitrogen, #130200). Areas of demyelination were defined as complete loss of myelin in PLP-stained sections. Stage of white matter lesion (active, mixed active/inactive, and inactive) was determined using established criteria[14]. For each section, masks were drawn to delineate lesional and non-lesional white matter areas, and were subsequently superimposed to adjacent sections to allow quantification of the expression of other primary antibodies. In each region of interest, microglia-macrophage inflammation was quantified using a color-based extraction software (chromogen-positive pixels/$mm^2$), while BAPP-positive axons were quantified using a positive element detection software (number of positive elements/$mm^2$)[15]. Sections were also impregnated with Palmgren's silver to demonstrate axons, and axonal density were quantified in the non-lesional anterior and lateral cortico-spinal, and in dorsal columns as described elsewhere[16].

***Statistical analysis***
Statistical analyses were performed using SPSS version 26 (SPSS, Chicago, IL). Differences in population characteristics between genotype groups were tested using Mann-Whitney or Crosstab Chi-square, as appropriate for variable distribution. To correctly analyze multiple observations per each case, generalized estimating equations were built to compare CD68, BAPP, tract areas and axonal counts (set as dependent variable) between TT and AT/AA *HIF1A* genotypes (set as independent variable), accounting for intra-subject variability. CD68 was added as covariate to the model if significantly different between genotypes, given the known correlation with BAPP[17], and cord level was included to correct analyses on tract area and axonal count. Pairwise comparison between estimated means was performed with Wald Chi-square, which is reported with its p-value. Missing values were treated as randomly missing values.

**Neurofilament levels**
***Plasma NFL***
Plasma samples for the study on NFL had been collected and frozen at the Karolinska Institutet and belonged to the IMSE cohort, collected to perform post-marketing disease-modifying treatments monitoring[18]. Plasma NFL levels were determined using a sensitive immunoassay on the Simoa platform through a commercially available NF-Light kit and antibodies from UmanDiagnostics according to the manufacturer's instructions (Quanterix, Lexington, MA), as previously described[19]. Patients were naive or had been treated with glatiramer acetate or interferon and were sampled before the beginning of the first or a new DMT. To study the effect of *HIF1A* genotype on plasma NFL levels, starting from this dataset we extracted all the patients belonging to the SWE cohort used for the genetic analysis and that had been sampled before 20 years from disease onset, when they had still a RR-MS course and were not in a relapse.
When evaluating the response to a new DMT in terms of change in plasma NFL, starting from the original IMSE cohort described above, we included all the RR-MS patients within 20 years from onset that had been sampled longitudinally at baseline and after treatment start (12±6 months).

***CSF NFL***
CSF NFL samples had been collected at the Karolinska Institutet at the time of neurological assessment that led to the diagnosis of MS[20]. NFL levels in the CSF were measured using commercially available ELISA kits (Uman diagnostics, Umeå, Sweden) according to the manufacturers' instructions.

***NFL levels analysis***
All NFL levels were used in linear models in the form of a z-score, after rank-based inverse normal transformation[21]. Plasma NFL levels were adjusted for body mass index and age at sampling as suggested by recent literature[22]; moreover, disease duration at sampling and line of treatment (first versus second line) about to start were used as covariates. In the CSF analysis, the NFL levels were adjusted for disease duration and age at sampling. To assess the effect of treatment on NFL reduction over time, we used mixed-effects linear models, to account for intra-subject variation and adjusting for disease duration.

**Pharmacogenomic study of dimethyl fumarate**
We evaluated how the rs11621525 genotype affected the response to dimethyl fumarate (DMF), using the No Evidence of Disease Activity status[23] (NEDA-3) at two years from treatment start as outcome of disease activity. The NEDA-3 status was defined when all of the following criteria were satisfied over the 2-years follow-up: a) no new relapses; b) no new/enlarging or gadolinium enhancing lesions at brain MRI; c) no Expanded Disability Status Scale (EDSS) progression over the follow-up. EDSS progression was defined as follows: a) an increase of at least 1.5 points for patients with baseline EDSS = 0; b) an increase of at least 1.0 point for baseline EDSS between 1.0 and 5.5; c) increase of at least 0.5 point for baseline EDSS ≥ 6.0.
From the cohort of patients with available whole-genome imputed genetic data, that are part of the cohort available at the Laboratory of Human Genetics of Neurological Disorders at San Raffaele Scientific Institute described above, we

identified 138 patients that fulfilled the following inclusion criteria: 1) diagnosis of RR-MS; 2) patients not previously treated with second line DMT or other immunosuppressive drugs; 3) continuous DMF treatment for at least 2-years or shift to other DMT for lack of efficacy of DMF; 4) availability of clinical information to define the NEDA-3 status at two-years from DMF start. Patients who discontinued DMF treatment for side effects within the first 2 years, with no evidence of disease activity under treatment, were not included in the analysis given that the NEDA-3 status could not be assessed at 2-year follow-up. Logistic regression was used to assess the effect of rs11621525 genotype (as predictor) on the NEDA-3 status at 2-years from treatment start (as outcome), adjusting for age at disease onset and sex.

**Ethics committees approval**

Patients participating to this study had given written informed consent and the studies had been approved locally by the respective ethical committee. Local approval numbers are as follows. San Raffaele Scientific Institute: 1013/DG, 274/DG, 397/ER/mm, 851/DG, 20/int/2019. Karolinska Institutet: 02-548, 2006/845-31/1, 2009/2107-31/2. 2011/641-31/4, 02-548, 2015/2235-32/4, 2015/2236-32, 2015/2260-32/2, 2016/1167-32, 2016/1168-32, 2016/1169-32, 2017/1347-32, 2017/1348-32, 2017/1349-32, 2017/1350-32, 2017/1426-32. Oxford Neuroinflammation Center: REC-08/MRE09/31+5.

**SUPPLEMENTARY FIGURES**

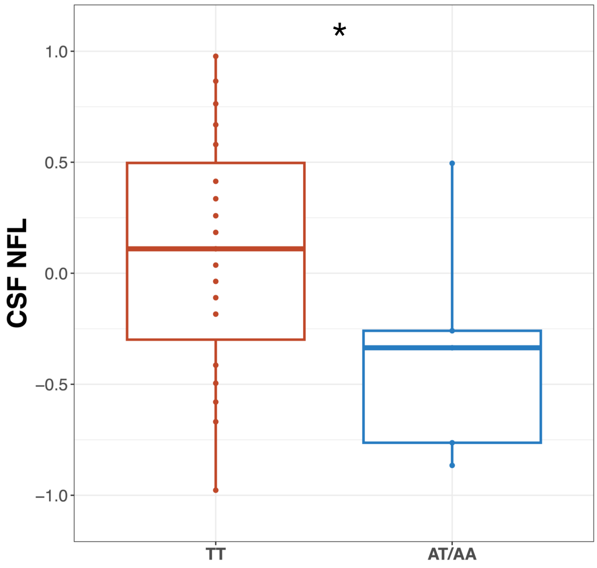

***eFigure 1. CSF NFL levels in CIS.***
On the x-axis the genotype for rs1951795 variant (tagging rs11621525) is shown; on the y-axis the z-score for normalized CSF NFL levels is reported. * = P < .05. CIS: Clinically Isolated Syndrome

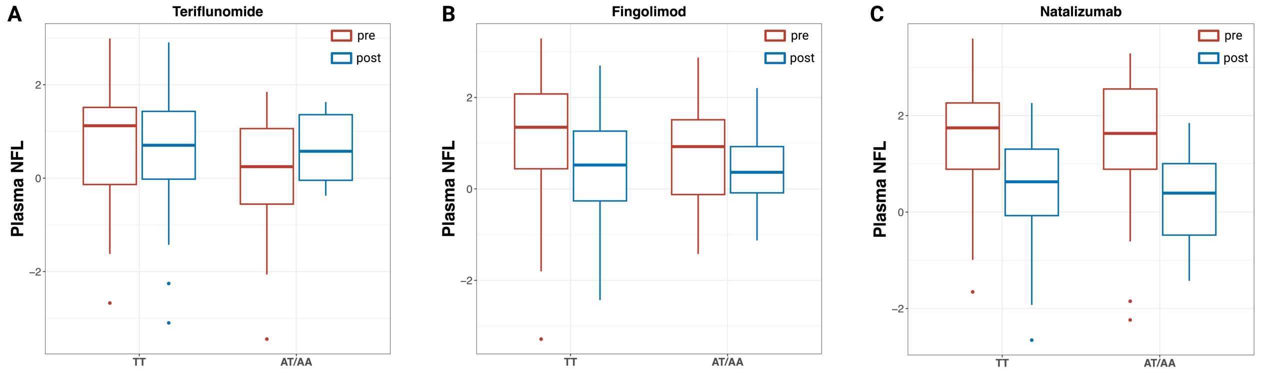

***eFigure 2. Effect of HIF1A genotype on plasma NFL levels change after the start of treatment with teriflunomide, fingolimod and natalizumab.***

On the x-axis the genotype for rs1951795 variant (tagging rs11621525) is shown; on the y-axis the z-score for normalized plasma NFL levels before (in red; "pre") and after (in blue; "post") treatment start for teriflunomide (panel A), fingolimod (panel B) and natalizumab (panel C). The reduction of plasma NFL levels after treatment start is clearly visible for Fingolimod and Natalizumab, and only in the TT group for Teriflunomide; no significant additional effect of the *HIF1A* rs1951795 SNP on the NFL levels emerged, despite what observed in the DMF group (see main manuscript, Figure 4C).

**SUPPLEMENTARY TABLES**

**eTable 1. List of the selected iron metabolism gene sets.**

In the leftmost column the name of the gene set is reported, as coded in the respective source (GO = Gene Ontology; HP = Human Phenotype; KEGG = Kyoto Encyclopedia of Genes and Genomes). N of genes = number of genes in the gene set.

| *Gene set* | *Source* | *N of genes* |
|---|---|---|
| *GOBP_IRON_ION_TRANSPORT* | GO | 79 |
| *GOBP_CELLULAR_IRON_ION_HOMEOSTASIS* | GO | 70 |
| *GOBP_RESPONSE_TO_IRON_ION* | GO | 28 |
| *GOBP_RESPONSE_TO_IRON_II_ION* | GO | 5 |
| *GOBP_RESPONSE_TO_IRON_III_ION* | GO | 5 |
| *GOBP_IRON_SULFUR_CLUSTER_ASSEMBLY* | GO | 24 |
| *GOBP_IRON_IMPORT_INTO_CELL* | GO | 11 |
| *GOBP_IRON_ION_TRANSMEMBRANE_TRANSPORT* | GO | 19 |
| *GOBP_REGULATION_OF_IRON_ION_TRANSPORT* | GO | 9 |
| *GOBP_REGULATION_OF_IRON_ION_TRANSMEMBRANE_TRANSPORT* | GO | 6 |
| *GOBP_IRON_ION_HOMEOSTASIS* | GO | 86 |
| *GOBP_MULTICELLULAR_ORGANISMAL_IRON_ION_HOMEOSTASIS* | GO | 7 |
| *GOBP_CELLULAR_RESPONSE_TO_IRON_ION* | GO | 9 |
| *GOBP_PROTEIN_MATURATION_BY_IRON_SULFUR_CLUSTER_TRANSFER* | GO | 16 |
| *GOBP_SEQUESTERING_OF_IRON_ION* | GO | 7 |
| *GOBP_IRON_ION_IMPORT_ACROSS_PLASMA_MEMBRANE* | GO | 6 |
| *GOBP_IRON_COORDINATION_ENTITY_TRANSPORT* | GO | 14 |
| *GOMF_IRON_ION_TRANSMEMBRANE_TRANSPORTER_ACTIVITY* | GO | 10 |
| *GOMF_IRON_ION_BINDING* | GO | 150 |
| *GOMF_FERROUS_IRON_BINDING* | GO | 26 |
| *GOMF_FERRIC_IRON_BINDING* | GO | 10 |
| *HP_ABNORMALITY_OF_IRON_HOMEOSTASIS* | HP | 19 |
| *HP_IRON_ACCUMULATION_IN_BRAIN* | HP | 9 |
| *KEGG_FERROPTOSIS* | KEGG | 41 |

**eTable 2. List of the mapped genes in iron metabolism included in the genetic analysis.**
For each of the 334 included genes, chromosome (CHR), positional information of the gene in base-pairs according to GRCh37/hg19 reference genome (START, END) and gene symbol (GENE) according to HUGO Gene Nomenclature Committee are reported. The final region included in the analysis (START-2kb and END+2kb) is also shown, representing the gene body with a 2 kilobase (kb) window at end. See full text for details.

| *CHR* | *START* | *END* | *GENE* | *START-2kb* | *END+2kb* |
|---|---|---|---|---|---|
| *1* | 11994723 | 12035599 | PLOD1 | 11992723 | 12037599 |
| *1* | 17312452 | 17338423 | ATP13A2 | 17310452 | 17340423 |
| *1* | 22963117 | 22966175 | C1QA | 22961117 | 22968175 |
| *1* | 43212005 | 43232755 | P3H1 | 43210005 | 43234755 |
| *1* | 44440601 | 44443972 | ATP6V0B | 44438601 | 44445972 |
| *1* | 44462154 | 44483012 | SLC6A9 | 44460154 | 44485012 |
| *1* | 47264669 | 47285021 | CYP4B1 | 47262669 | 47287021 |
| *1* | 47308766 | 47366147 | CYP4Z2P | 47306766 | 47368147 |
| *1* | 47394845 | 47407156 | CYP4A11 | 47392845 | 47409156 |
| *1* | 47489239 | 47516423 | CYP4X1 | 47487239 | 47518423 |
| *1* | 47533159 | 47583992 | CYP4Z1 | 47531159 | 47585992 |
| *1* | 47603106 | 47614526 | CYP4A22 | 47601106 | 47616526 |
| *1* | 60358979 | 60392423 | CYP2J2 | 60356979 | 60394423 |
| *1* | 94352589 | 94375012 | GCLM | 94350589 | 94377012 |
| *1* | 109648572 | 109656479 | C1orf194 | 109646572 | 109658479 |
| *1* | 145413190 | 145417545 | HJV | 145411190 | 145419545 |
| *1* | 155259083 | 155271225 | PKLR | 155257083 | 155273225 |
| *1* | 198492351 | 198510075 | ATP6V1G3 | 198490351 | 198512075 |
| *1* | 213031596 | 213072705 | FLVCR1 | 213029596 | 213074705 |
| *1* | 220087605 | 220101993 | SLC30A10 | 220085605 | 220103993 |
| *1* | 228353428 | 228369958 | IBA57 | 228351428 | 228371958 |
| *1* | 231499496 | 231560790 | EGLN1 | 231497496 | 231562790 |
| *1* | 235530727 | 235612280 | TBCE | 235528727 | 235614280 |
| *1* | 242158791 | 242162385 | MAP1LC3C | 242156791 | 242164385 |
| *2* | 3501689 | 3523350 | ADI1 | 3499689 | 3525350 |
| *2* | 10861774 | 10925236 | ATP6V1C2 | 10859774 | 10927236 |
| *2* | 31557187 | 31637611 | XDH | 31555187 | 31639611 |
| *2* | 38294745 | 38303323 | CYP1B1 | 38292745 | 38305323 |
| *2* | 42994228 | 43019751 | HAAO | 42992228 | 43021751 |
| *2* | 46524540 | 46613842 | EPAS1 | 46522540 | 46615842 |
| *2* | 46755024 | 46769141 | ATP6V1E2 | 46753024 | 46771141 |
| *2* | 47168312 | 47303275 | TTC7A | 47166312 | 47305275 |
| *2* | 69623244 | 69664760 | NFU1 | 69621244 | 69666760 |
| *2* | 70314584 | 70316334 | PCBP1 | 70312584 | 70318334 |
| *2* | 71162997 | 71192561 | ATP6V1B1 | 71160997 | 71194561 |
| *2* | 72356366 | 72374991 | CYP26B1 | 72354366 | 72376991 |
| *2* | 86668583 | 86719839 | KDM3A | 86666583 | 86721839 |
| *2* | 96931883 | 96939900 | CIAO1 | 96929883 | 96941900 |
| *2* | 119981383 | 120023227 | STEAP3 | 119979383 | 120025227 |
| *2* | 127941411 | 127963343 | CYP27C1 | 127939411 | 127965343 |
| *2* | 131095815 | 131099922 | CCDC115 | 131093815 | 131101922 |
| *2* | 172378865 | 172414643 | CYBRD1 | 172376865 | 172416643 |
| *2* | 190425315 | 190445537 | SLC40A1 | 190423315 | 190447537 |
| *2* | 200793633 | 200820459 | TYW5 | 200791633 | 200822459 |
| *2* | 201450730 | 201536217 | AOX1 | 201448730 | 201538217 |
| *2* | 204103163 | 204170563 | CYP20A1 | 204101163 | 204172563 |
| *2* | 219246751 | 219261617 | SLC11A1 | 219244751 | 219263617 |
| *2* | 219524378 | 219528166 | BCS1L | 219522378 | 219530166 |
| *2* | 219646471 | 219680016 | CYP27A1 | 219644471 | 219682016 |
| *2* | 220074487 | 220085174 | RP11-803J6.1, ABCB6 | 220072487 | 220087174 |
| *2* | 223725731 | 223808119 | ACSL3 | 223723731 | 223810119 |
| *2* | 239072632 | 239077515 | ERFE | 239070632 | 239079515 |
| *3* | 3168599 | 3190706 | TRNT1 | 3166599 | 3192706 |
| *3* | 11314009 | 11599139 | ATG7 | 11312009 | 11601139 |

| | | | | | |
|---|---|---|---|---|---|
| *3* | 42913683 | 42917633 | CYP8B1 | 42911683 | 42919633 |
| *3* | 46477495 | 46506598 | LTF | 46475495 | 46508598 |
| *3* | 48601505 | 48632593 | COL7A1 | 48599505 | 48634593 |
| *3* | 49027340 | 49044581 | P4HTM | 49025340 | 49046581 |
| *3* | 50355220 | 50360281 | HYAL2 | 50353220 | 50362281 |
| *3* | 71003864 | 71180092 | FOXP1 | 71001864 | 71182092 |
| *3* | 113465865 | 113530905 | ATP6V1A | 113463865 | 113532905 |
| *3* | 133464799 | 133497850 | TF | 133462799 | 133499850 |
| *3* | 145787227 | 145879282 | PLOD2 | 145785227 | 145881282 |
| *3* | 148890289 | 148939832 | CP | 148888289 | 148941832 |
| *3* | 184428154 | 184429836 | MAGEF1 | 184426154 | 184431836 |
| *3* | 186383797 | 186396023 | HRG | 186381797 | 186398023 |
| *3* | 189674516 | 189838908 | P3H2 | 189672516 | 189840908 |
| *3* | 195776154 | 195809032 | TFRC | 195774154 | 195811032 |
| *3* | 196728611 | 196756687 | MELTF | 196726611 | 196758687 |
| *4* | 15606006 | 15657035 | FBXL5 | 15604006 | 15659035 |
| *4* | 76781025 | 76823681 | PPEF2 | 76779025 | 76825681 |
| *4* | 83550689 | 83720010 | SCD5 | 83548689 | 83722010 |
| *4* | 89011415 | 89080011 | ABCG2 | 89009415 | 89082011 |
| *4* | 90645249 | 90759447 | SNCA | 90643249 | 90761447 |
| *4* | 103182820 | 103266655 | SLC39A8 | 103180820 | 103268655 |
| *4* | 103998781 | 104021024 | BDH2 | 103996781 | 104023024 |
| *4* | 106067841 | 106200960 | TET2 | 106065841 | 106202960 |
| *4* | 108852716 | 108874613 | CYP2U1 | 108850716 | 108876613 |
| *4* | 129190391 | 129209984 | PGRMC2 | 129188391 | 129211984 |
| *4* | 139085247 | 139163503 | SLC7A11 | 139083247 | 139165503 |
| *4* | 146019155 | 146050676 | ABCE1 | 146017155 | 146052676 |
| *4* | 166248817 | 166264314 | MSMO1 | 166246817 | 166266314 |
| *4* | 185676748 | 185747215 | ACSL1 | 185674748 | 185749215 |
| *4* | 187112673 | 187134617 | CYP4V2 | 187110673 | 187136617 |
| *5* | 1392904 | 1445543 | SLC6A3 | 1390904 | 1447543 |
| *5* | 68462836 | 68474070 | CCNB1 | 68460836 | 68476070 |
| *5* | 94799598 | 94890709 | TTC37 | 94797598 | 94892709 |
| *5* | 115140429 | 115152405 | CDO1 | 115138429 | 115154405 |
| *5* | 121187649 | 121188523 | FTMT | 121185649 | 121190523 |
| *5* | 131285666 | 131347355 | ACSL6 | 131283666 | 131349355 |
| *5* | 131584600 | 131631008 | P4HA2 | 131582600 | 131633008 |
| *5* | 133492081 | 133512724 | SKP1 | 133490081 | 133514724 |
| *5* | 137890570 | 137911318 | HSPA9 | 137888570 | 137913318 |
| *5* | 141488323 | 141534008 | NDFIP1 | 141486323 | 141536008 |
| *5* | 154198051 | 154230213 | FAXDC2 | 154196051 | 154232213 |
| *5* | 172410762 | 172461900 | ATP6V0E1 | 172408762 | 172463900 |
| *6* | 5186833 | 5261172 | LYRM4 | 5184833 | 5263172 |
| *6* | 7727010 | 7881961 | BMP6 | 7725010 | 7883961 |
| *6* | 10396915 | 10415470 | TFAP2A | 10394915 | 10417470 |
| *6* | 26087508 | 26095469 | HFE | 26085508 | 26097469 |
| *6* | 31512227 | 31514625 | ATP6V1G2 | 31510227 | 31516625 |
| *6* | 31926580 | 31937532 | SKIV2L | 31924580 | 31939532 |
| *6* | 31973358 | 31976712 | CYP21A1P, CYP21A | 31971358 | 31978712 |
| *6* | 32006092 | 32009447 | CYP21A2 | 32004092 | 32011447 |
| *6* | 38136226 | 38607924 | BTBD9 | 38134226 | 38609924 |
| *6* | 46517444 | 46620523 | CYP39A1 | 46515444 | 46622523 |
| *6* | 53362139 | 53409927 | GCLC | 53360139 | 53411927 |
| *6* | 106632351 | 106773695 | ATG5 | 106630351 | 106775695 |
| *7* | 1022834 | 1029276 | CYP2W1 | 1020834 | 1031276 |
| *7* | 6061877 | 6098860 | EIF2AK1 | 6059877 | 6100860 |
| *7* | 15239942 | 15601640 | AGMO | 15237942 | 15603640 |
| *7* | 39606002 | 39612480 | YAE1 | 39604002 | 39614480 |
| *7* | 87834431 | 87849399 | SRI | 87832431 | 87851399 |
| *7* | 87905743 | 87936228 | STEAP4 | 87903743 | 87938228 |
| *7* | 89783688 | 89794141 | STEAP1 | 89781688 | 89796141 |

| | | | | | |
|---|---|---|---|---|---|
| *7* | 89840999 | 89866992 | STEAP2 | 89838999 | 89868992 |
| *7* | 91741462 | 91763840 | CYP51A1 | 91739462 | 91765840 |
| *7* | 97736196 | 97838944 | LMTK2 | 97734196 | 97840944 |
| *7* | 99282301 | 99332819 | CYP3A7-CYP3A51P, CYP3A7 | 99280301 | 99334819 |
| *7* | 99354582 | 99381811 | CYP3A4 | 99352582 | 99383811 |
| *7* | 99425635 | 99464173 | CYP3A43 | 99423635 | 99466173 |
| *7* | 100218038 | 100239201 | TFR2 | 100216038 | 100241201 |
| *7* | 100849257 | 100861011 | PLOD3 | 100847257 | 100863011 |
| *7* | 128502856 | 128505903 | ATP6V1F | 128500856 | 128507903 |
| *7* | 138391038 | 138458782 | ATP6V0A4 | 138389038 | 138460782 |
| *7* | 139528951 | 139720125 | TBXAS1 | 139526951 | 139722125 |
| *7* | 139784545 | 139876741 | KDM7A | 139782545 | 139878741 |
| *7* | 148395932 | 148498202 | CUL1 | 148393932 | 148500202 |
| *7* | 149570056 | 149577787 | ATP6V0E2 | 149568056 | 149579787 |
| *7* | 150745378 | 150749843 | ASIC3 | 150743378 | 150751843 |
| *8* | 20054703 | 20079207 | ATP6V1B2 | 20052703 | 20081207 |
| *8* | 22225049 | 22280249 | SLC39A14 | 22223049 | 22282249 |
| *8* | 23386362 | 23430063 | SLC25A37 | 23384362 | 23432063 |
| *8* | 27727398 | 27850369 | SCARA5 | 27725398 | 27852369 |
| *8* | 42249278 | 42263455 | VDAC3 | 42247278 | 42265455 |
| *8* | 54628102 | 54755871 | ATP6V1H | 54626102 | 54757871 |
| *8* | 59402736 | 59412720 | CYP7A1 | 59400736 | 59414720 |
| *8* | 65508528 | 65711348 | CYP7B1 | 65506528 | 65713348 |
| *8* | 87111138 | 87166454 | ATP6V0D2 | 87109138 | 87168454 |
| *8* | 104033247 | 104085285 | ATP6V1C1 | 104031247 | 104087285 |
| *8* | 128748314 | 128753680 | MYC | 128746314 | 128755680 |
| *8* | 143953772 | 143961236 | CYP11B1 | 143951772 | 143963236 |
| *8* | 143991974 | 143999259 | CYP11B2 | 143989974 | 144001259 |
| *9* | 32384600 | 32450832 | ACO1 | 32382600 | 32452832 |
| *9* | 71650478 | 71693993 | FXN | 71648478 | 71695993 |
| *9* | 79792360 | 80032399 | VPS13A | 79790360 | 80034399 |
| *9* | 88879462 | 88897490 | ISCA1 | 88877462 | 88899490 |
| *9* | 96338908 | 96441869 | PHF2 | 96336908 | 96443869 |
| *9* | 116148591 | 116163618 | ALAD | 116146591 | 116165618 |
| *9* | 117349993 | 117361152 | ATP6V1G1 | 117347993 | 117363152 |
| *9* | 130911731 | 130915734 | LCN2 | 130909731 | 130917734 |
| *9* | 139257440 | 139268133 | CARD9 | 139255440 | 139270133 |
| *9* | 140100118 | 140113813 | NDOR1 | 140098118 | 140115813 |
| *10* | 13319795 | 13342130 | PHYH | 13317795 | 13344130 |
| *10* | 45869623 | 45941567 | ALOX5 | 45867623 | 45943567 |
| *10* | 48413091 | 48416853 | GDF2 | 48411091 | 48418853 |
| *10* | 51565107 | 51590734 | NCOA4 | 51563107 | 51592734 |
| *10* | 70320116 | 70454239 | TET1 | 70318116 | 70456239 |
| *10* | 74766979 | 74856732 | P4HA1 | 74764979 | 74858732 |
| *10* | 76969911 | 76991207 | VDAC2 | 76967911 | 76993207 |
| *10* | 90965693 | 90967071 | CH25H | 90963693 | 90969071 |
| *10* | 94821020 | 94828454 | CYP26C1 | 94819020 | 94830454 |
| *10* | 94833646 | 94837641 | CYP26A1 | 94831646 | 94839641 |
| *10* | 96522462 | 96612671 | CYP2C19 | 96520462 | 96614671 |
| *10* | 96698414 | 96749148 | CYP2C9 | 96696414 | 96751148 |
| *10* | 96796528 | 96829254 | CYP2C8 | 96794528 | 96831254 |
| *10* | 99218080 | 99258366 | MMS19 | 99216080 | 99260366 |
| *10* | 101370274 | 101380221 | SLC25A28 | 101368274 | 101382221 |
| *10* | 102106771 | 102124588 | SCD | 102104771 | 102126588 |
| *10* | 102295640 | 102313681 | HIF1AN | 102293640 | 102315681 |
| *10* | 104590287 | 104597290 | CYP17A1 | 104588287 | 104599290 |
| *10* | 114135955 | 114188138 | ACSL5 | 114133955 | 114190138 |
| *10* | 131934638 | 131977932 | GLRX3 | 131932638 | 131979932 |
| *10* | 135340866 | 135352620 | CYP2E1 | 135338866 | 135354620 |
| *11* | 568088 | 568198 | MIR210 | 566088 | 570198 |

| | | | | | |
|---|---|---|---|---|---|
| *11* | 2185158 | 2193035 | TH | 2183158 | 2195035 |
| *11* | 5246695 | 5248301 | HBB | 5244695 | 5250301 |
| *11* | 6452267 | 6462254 | HPX | 6450267 | 6464254 |
| *11* | 14899555 | 14913751 | CYP2R1 | 14897555 | 14915751 |
| *11* | 18042083 | 18062335 | TPH1 | 18040083 | 18064335 |
| *11* | 27062508 | 27149354 | BBOX1 | 27060508 | 27151354 |
| *11* | 31391376 | 31454382 | DNAJC24 | 31389376 | 31456382 |
| *11* | 34937676 | 35017675 | PDHXPDX1 | 34935676 | 35019675 |
| *11* | 43902356 | 43941825 | ALKBH3 | 43900356 | 43943825 |
| *11* | 46698624 | 46722215 | ARHGAP1 | 46696624 | 46724215 |
| *11* | 61731756 | 61735132 | FTH1 | 61729756 | 61737132 |
| *11* | 62623483 | 62656355 | SLC3A2 | 62621483 | 62658355 |
| *11* | 67806461 | 67818366 | TCIRG1 | 67804461 | 67820366 |
| *11* | 69455872 | 69469242 | CCND1 | 69453872 | 69471242 |
| *11* | 69480331 | 69490165 | LTO1 | 69478331 | 69492165 |
| *11* | 73977701 | 74022699 | P4HA3 | 73975701 | 74024699 |
| *11* | 85668213 | 85780139 | PICALM | 85666213 | 85782139 |
| *11* | 93754377 | 93847374 | HEPHL1 | 93752377 | 93849374 |
| *11* | 107373452 | 107436461 | ALKBH8 | 107371452 | 107438461 |
| *11* | 110300660 | 110335608 | FDX1 | 110298660 | 110337608 |
| *11* | 111895537 | 111935002 | DLAT | 111893537 | 111937002 |
| *11* | 113280316 | 113346001 | DRD2 | 113278316 | 113348001 |
| *11* | 119531702 | 119599435 | NECTIN1 | 119529702 | 119601435 |
| *11* | 121163387 | 121184119 | SC5D | 121161387 | 121186119 |
| *12* | 7085346 | 7125842 | LPCAT3 | 7083346 | 7127842 |
| *12* | 27849427 | 27850566 | REP15 | 27847427 | 27852566 |
| *12* | 46576840 | 46663208 | SLC38A1, SAT1 | 46574840 | 46665208 |
| *12* | 46751970 | 46766645 | SLC38A2, SAT2 | 46749970 | 46768645 |
| *12* | 48166966 | 48176536 | SLC48A1 | 48164966 | 48178536 |
| *12* | 51379774 | 51422058 | SLC11A2 | 51377774 | 51424058 |
| *12* | 53845885 | 53874946 | PCBP2 | 53843885 | 53876946 |
| *12* | 58156116 | 58160976 | CYP27B1 | 58154116 | 58162976 |
| *12* | 67663060 | 67708388 | CAND1 | 67661060 | 67710388 |
| *12* | 68548549 | 68553521 | IFNG | 68546549 | 68555521 |
| *12* | 72332625 | 72426221 | TPH2 | 72330625 | 72428221 |
| *12* | 103232103 | 103311381 | PAH | 103230103 | 103313381 |
| *12* | 108956293 | 108963160 | ISCU | 108954293 | 108965160 |
| *12* | 109525992 | 109531293 | ALKBH2 | 109523992 | 109533293 |
| *12* | 116997185 | 117014425 | MAP1LC3B2 | 116995185 | 117016425 |
| *12* | 123459353 | 123464588 | OGFOD2 | 123457353 | 123466588 |
| *12* | 124196864 | 124246301 | ATP6V0A2 | 124194864 | 124248301 |
| *13* | 28494167 | 28500451 | PDX1 | 28492167 | 28502451 |
| *14* | 23815526 | 23821660 | SLC22A17 | 23813526 | 23823660 |
| *14* | 32030590 | 32330429 | NUBPL | 32028590 | 32332429 |
| *14* | 34393420 | 34420284 | EGLN3 | 34391420 | 34422284 |
| *14* | 62162118 | 62214977 | HIF1A | 62160118 | 62216977 |
| *14* | 67804580 | 67826720 | ATP6V1D | 67802580 | 67828720 |
| *14* | 74960422 | 74962271 | ISCA2 | 74958422 | 74964271 |
| *14* | 76044939 | 76114512 | FLVCR2 | 76042939 | 76116512 |
| *14* | 78138748 | 78174356 | ALKBH1 | 78136748 | 78176356 |
| *14* | 96001322 | 96011055 | GLRX5 | 95999322 | 96013055 |
| *14* | 100150754 | 100193638 | CYP46A1 | 100148754 | 100195638 |
| *15* | 43489425 | 43513323 | EPB42 | 43487425 | 43515323 |
| *15* | 45003684 | 45010357 | B2M | 45001684 | 45012357 |
| *15* | 51500253 | 51630795 | CYP19A1 | 51498253 | 51632795 |
| *15* | 64364760 | 64386207 | CIAO2A | 64362760 | 64388207 |
| *15* | 69307033 | 69349501 | NOX5 | 69305033 | 69351501 |
| *15* | 69706626 | 69740764 | KIF23 | 69704626 | 69742764 |
| *15* | 73344824 | 73597547 | NEO1 | 73342824 | 73599547 |
| *15* | 74630102 | 74660081 | CYP11A1 | 74628102 | 74662081 |
| *15* | 75011882 | 75017877 | CYP1A1 | 75009882 | 75019877 |

| | | | | | |
|---|---|---|---|---|---|
| *15* | 75041183 | 75048941 | CYP1A2 | 75039183 | 75050941 |
| *15* | 78730517 | 78793798 | IREB2 | 78728517 | 78795798 |
| *16* | 202853 | 204504 | HBZ | 200853 | 206504 |
| *16* | 222845 | 223709 | HBA2 | 220845 | 225709 |
| *16* | 226678 | 227520 | HBA1 | 224678 | 229520 |
| *16* | 230332 | 231178 | HBQ1 | 228332 | 233178 |
| *16* | 779768 | 790997 | CIAO3 | 777768 | 792997 |
| *16* | 1832932 | 1839192 | NUBP2 | 1830932 | 1841192 |
| *16* | 2563726 | 2570224 | ATP6V0C | 2561726 | 2572224 |
| *16* | 4526340 | 4560348 | HMOX2 | 4524340 | 4562348 |
| *16* | 10837697 | 10863208 | NUBP1 | 10835697 | 10865208 |
| *16* | 29464913 | 29466285 | BOLA2 | 29462913 | 29468285 |
| *16* | 30204255 | 30205627 | BOLA2B | 30202255 | 30207627 |
| *16* | 53737874 | 54148379 | FTO | 53735874 | 54150379 |
| *16* | 56485423 | 56511407 | OGFOD1 | 56483423 | 56513407 |
| *16* | 57462086 | 57481369 | CIAPIN1 | 57460086 | 57483369 |
| *16* | 66965957 | 66968326 | CIAO2B | 66963957 | 66970326 |
| *16* | 67471916 | 67515089 | ATP6V0D1 | 67469916 | 67517089 |
| *16* | 74746855 | 74808729 | FA2H | 74744855 | 74810729 |
| *16* | 87425800 | 87438380 | MAP1LC3B | 87423800 | 87440380 |
| *17* | 4534213 | 4544971 | ALOX15 | 4532213 | 4546971 |
| *17* | 6899383 | 6914055 | ALOX12 | 6897383 | 6916055 |
| *17* | 7529555 | 7531194 | SAT2 | 7527555 | 7533194 |
| *17* | 7571719 | 7590868 | TP53 | 7569719 | 7592868 |
| *17* | 7942357 | 7952451 | ALOX15B | 7940357 | 7954451 |
| *17* | 7975953 | 7991021 | ALOX12B | 7973953 | 7993021 |
| *17* | 7999217 | 8022234 | ALOXE3 | 7997217 | 8024234 |
| *17* | 26684686 | 26689089 | TMEM199 | 26682686 | 26691089 |
| *17* | 26721660 | 26733230 | SLC46A1 | 26719660 | 26735230 |
| *17* | 36886509 | 36891858 | CISD3 | 36884509 | 36893858 |
| *17* | 40610861 | 40674597 | ATP6V0A1 | 40608861 | 40676597 |
| *17* | 40962149 | 40976310 | BECN1 | 40960149 | 40978310 |
| *17* | 57697049 | 57774317 | CLTC | 57695049 | 57776317 |
| *17* | 74523429 | 74533987 | CYGB | 74521429 | 74535987 |
| *17* | 74708913 | 74722881 | JMJD6 | 74706913 | 74724881 |
| *17* | 80347085 | 80376513 | OGFOD3 | 80345085 | 80378513 |
| *18* | 48556582 | 48611411 | SMAD4 | 48554582 | 48613411 |
| *18* | 55212072 | 55253969 | FECH | 55210072 | 55255969 |
| *18* | 60790578 | 60986613 | BCL2 | 60788578 | 60988613 |
| *19* | 1104648 | 1106787 | GPX4 | 1102648 | 1108787 |
| *19* | 3490818 | 3500621 | DOHH | 3488818 | 3502621 |
| *19* | 7587495 | 7598895 | MCOLN1 | 7585495 | 7600895 |
| *19* | 8455204 | 8469317 | RAB11B | 8453204 | 8471317 |
| *19* | 10828728 | 10942586 | DNM2 | 10826728 | 10944586 |
| *19* | 11685474 | 11689801 | ACP5 | 11683474 | 11691801 |
| *19* | 13049413 | 13055304 | CALR | 13047413 | 13057304 |
| *19* | 15619335 | 15663128 | CYP4F22 | 15617335 | 15665128 |
| *19* | 15726028 | 15740447 | CYP4F8 | 15724028 | 15742447 |
| *19* | 15751706 | 15771570 | CYP4F3 | 15749706 | 15773570 |
| *19* | 15783827 | 15807984 | CYP4F12 | 15781827 | 15809984 |
| *19* | 15988833 | 16008884 | CYP4F2 | 15986833 | 16010884 |
| *19* | 16023179 | 16045676 | CYP4F11 | 16021179 | 16047676 |
| *19* | 35773409 | 35776045 | HAMP | 35771409 | 35778045 |
| *19* | 41305333 | 41314346 | EGLN2 | 41303333 | 41316346 |
| *19* | 41349442 | 41356352 | CYP2A6 | 41347442 | 41358352 |
| *19* | 41381343 | 41388657 | CYP2A7 | 41379343 | 41390657 |
| *19* | 41396730 | 41406413 | CYP2A6 | 41394730 | 41408413 |
| *19* | 41497203 | 41524301 | CYP2B6 | 41495203 | 41526301 |
| *19* | 41594355 | 41602100 | CYP2A13 | 41592355 | 41604100 |
| *19* | 41620352 | 41634281 | CYP2F1 | 41618352 | 41636281 |
| *19* | 41699114 | 41713444 | CYP2S1 | 41697114 | 41715444 |

| | | | | | |
|---|---|---|---|---|---|
| *19* | 44010870 | 44031396 | ETHE1 | 44008870 | 44033396 |
| *19* | 49468565 | 49470136 | FTL | 49466565 | 49472136 |
| *19* | 54926604 | 54947899 | TTYH1 | 54924604 | 54949899 |
| *20* | 3869741 | 3904502 | PANK2 | 3867741 | 3906502 |
| *20* | 4666796 | 4682234 | PRNP | 4664796 | 4684234 |
| *20* | 6748744 | 6760910 | BMP2 | 6746744 | 6762910 |
| *20* | 33146500 | 33148149 | MAP1LC3A | 33144500 | 33150149 |
| *20* | 33516235 | 33543601 | GSS | 33514235 | 33545601 |
| *20* | 34256609 | 34287287 | NFS1 | 34254609 | 34289287 |
| *20* | 48120410 | 48184707 | PTGIS | 48118410 | 48186707 |
| *20* | 52769987 | 52790516 | CYP24A1 | 52767987 | 52792516 |
| *21* | 33031934 | 33041243 | SOD1 | 33029934 | 33043243 |
| *21* | 43782390 | 43786644 | TFF1 | 43780390 | 43788644 |
| *22* | 18074902 | 18111588 | ATP6V1E1 | 18072902 | 18113588 |
| *22* | 29138042 | 29153496 | HSCB | 29136042 | 29155496 |
| *22* | 35777059 | 35790207 | HMOX1 | 35775059 | 35792207 |
| *22* | 37461478 | 37499693 | TMPRSS6 | 37459478 | 37501693 |
| *22* | 38507501 | 38577761 | PLA2G6 | 38505501 | 38579761 |
| *22* | 41865128 | 41924993 | ACO2 | 41863128 | 41926993 |
| *22* | 42522500 | 42526883 | CYP2D6 | 42520500 | 42528883 |
| *22* | 42536213 | 42540575 | CYP2D7 | 42534213 | 42542575 |
| *22* | 50925212 | 50928750 | MIOX | 50923212 | 50930750 |
| *X* | 18709044 | 18846034 | PPEF1 | 18707044 | 18848034 |
| *X* | 23801274 | 23804327 | SAT1 | 23799274 | 23806327 |
| *X* | 31089357 | 31090170 | FTHL17 | 31087357 | 31092170 |
| *X* | 37639269 | 37672714 | CYBB | 37637269 | 37674714 |
| *X* | 48932091 | 48937564 | WDR45 | 48930091 | 48939564 |
| *X* | 53963112 | 54071569 | PHF8 | 53961112 | 54073569 |
| *X* | 55035487 | 55057497 | ALAS2 | 55033487 | 55059497 |
| *X* | 65382432 | 65487230 | HEPH | 65380432 | 65489230 |
| *X* | 74273006 | 74376175 | ABCB7 | 74271006 | 74378175 |
| *X* | 77166193 | 77305892 | ATP7A | 77164193 | 77307892 |
| *X* | 108884563 | 108976621 | ACSL4 | 108882563 | 108978621 |
| *X* | 135044230 | 135056134 | MMGT1 | 135042230 | 135058134 |
| *X* | 153656977 | 153664862 | ATP6AP1 | 153654977 | 153666862 |
| *X* | 153759605 | 153775233 | G6PD | 153757605 | 153777233 |
| *X* | 154718672 | 154842622 | TMLHE | 154716672 | 154844622 |

**eTable 3. Results of the genetic association study in the ITA cohort.**

| *CHR* | *SNP* | *POS* | *A1* | *OR SP* | *95% CI* | *P-value* |
|---|---|---|---|---|---|---|
| 14 | rs11621525 | 62203056 | A | 0.57 | 0.44-0.72 | $3.30x10^{-6}$ |
| 14 | rs10873142 | 62203462 | C | 0.59 | 0.47-0.75 | $1.34x10^{-5}$ |
| 14 | rs4899057 | 62202942 | G | 0.59 | 0.47-0.75 | $1.34x10^{-5}$ |
| 14 | rs4902082 | 62212675 | C | 0.59 | 0.47-0.75 | $1.34x10^{-5}$ |
| 14 | rs12435848 | 62176891 | A | 0.61 | 0.49-0.77 | $2.11x10^{-5}$ |
| 14 | rs1951795 | 62171426 | A | 0.60 | 0.48-0.76 | $2.66x10^{-5}$ |
| 14 | rs12232182 | 62176220 | C | 0.65 | 0.53-0.82 | $1.50x10^{-4}$ |
| 14 | rs2301111 | 62200201 | G | 0.65 | 0.52-0.81 | $1.73x10^{-4}$ |
| 14 | rs7153817 | 62201500 | G | 0.65 | 0.52-0.81 | $1.73x10^{-4}$ |

CHR = chromosome; SNP = Single Nucleotide Polymorphism rsID; POS = position on chromosome (reference genome GRCh37/hg19); A1 = effect allele; OR SP= odds ratio of conversion to SP-MS for A1; 95% CI = 95% Confidence Interval for OR SP; P-value = nominal p-value from logistic regression as described in the Methods.

**eTable 4. Replication in the Swedish (SWE) cohort.**

| *CHR* | *SNP* | *BP* | *A1* | *NMISS* | *OR SP* | *95% CI* | *P* | *LD* |
|---|---|---|---|---|---|---|---|---|
| 14 | rs11621525 | 62203056 | A | 2056 | 0.80 | 0.67-0.95 | .012 | 1.00 |
| 14 | rs10873142 | 62203462 | C | 2042 | 0.79 | 0.67-0.94 | .0080 | 0.98 |
| 14 | rs4899057 | 62202942 | G | 2045 | 0.80 | 0.67-0.94 | .0087 | 0.98 |
| 14 | rs4902082 | 62212675 | C | 2032 | 0.80 | 0.67-0.95 | .0095 | 0.98 |
| 14 | rs1951795 | 62171426 | A | 2062 | 0.79 | 0.67-0.94 | .0079 | 0.98 |

CHR = chromosome; SNP = Single Nucleotide Polymorphism rsID; POS = position on chromosome (reference genome GRCh37/hg19); A1 = effect allele; NMISS = number of observations (=final number of subjects with non-missing genotype, phenotype or covariates); OR SP= odds ratio of conversion to SP-MS course for A1; 95% CI = 95% Confidence Interval for OR SP; P-value = nominal p-value from logistic regression; LD = $r^2$ indicating linkage disequilibrium (LD) with rs11621525.

**eTable 5. Characteristics of the cohort included in the post-mortem pathology study.**

| | ***TT*** ***108 samples from 36 cases*** | ***AT/AA*** ***51 samples from 17 cases*** | ***p-value*** |
|---|---|---|---|
| ***Sex*** | F: 64% (23)<br>M: 36% (13) | F: 82% (14)<br>M: 18% (3) | n.s. |
| ***Course*** | SP-MS: 84% (27)<br>PP-MS: 16% (5) | SP-MS: 93% (13)<br>PP-MS: 7% (1) | n.s. |
| ***HLA-DRB1*15:01+*** | 53.3% (16) | 47% (8) | n.s. |
| ***Brain weight (g)*** | 1174 (894-1380) | 1132 (931-1300) | n.s. |
| ***Post-mortem interval (h)*** | 16.69 (6-28) | 20.56 (7-38) | n.s. |
| ***Age at death (years)*** | 61.22 (40-92) | 65.24 (55-82) | n.s. |
| ***Disease duration (years)*** | 29.36 (12-56) | 31.19 (17-58) | n.s. |

For sex, course and HLA-DRB1*15:01 status, the number of patients (in brackets) and percentages are reported. For brain weight, post-mortem interval, age at death and disease duration mean value and range (in brackets) are reported. The p-value from chi-square test is reported for sex, course, and *HLA-DRB1*15:01* status. For the remaining, p-values from non-parametric Mann-Whitney test are reported. Number of subjects with missing information are as follows (TT group, AT/AA group). Course: 4, 3; *HLA-DRB1*15:01* status: 6, 0; brain weight: 1, 0; disease duration: 0, 1. Abbreviations: M = male; F = female; SP-MS = secondary progressive MS; PP-MS = primary progressive MS; *HLA-DRB1*15:01+* = positivity for *HLA-DRB1*15:01* allele; g = grams; h = hours; n.s. = not statistically significant.

**eTable 6. Effect of *HIF1A* genotype on non-lesional pathology.**

| | ***TT, 88 samples from 36 cases*** | | ***AT/AA, 40 samples from 17 cases*** | |
|---|---|---|---|---|
| | **Mean** | **95% Wald CI** | **Mean** | **95% Wald CI** |
| **ACST area ($mm^2$)** | 1.58 | 1.4 to 1.77 | 1.85 | 1.42 to 2.28 |
| **LCST area (mm2)** | 3.81 | 3.34 to 4.28 | 4.07 | 3.21 to 4.94 |
| **DC area ($mm^2$)** | 5.28 | 4.72 to 5.83 | 6.33 | 4.97 to 7.69 |
| **ACST total axons** | 29165 | 24264 to 34066 | 39437 | 28433 to 50442 |
| **LCST total axons** | 59600 | 47187 to 72013 | 62523 | 39918 to 85128 |
| **DC total axons** | 104098 | 90797 to 117398 | 128882 | 86711 to 171054 |

The table shows the notional tract cross-sectional areas (in $mm^2$) and total axonal counts in non-lesional anterior cortico-spinal tract (ACST), lateral cortico-spinal tract (LCST), and dorsal columns (DC) in *HIF1A*-TT and -AT/AA subgroups. Estimated mean values and 95% Wald confidence interval (CI) corrected for cord level are displayed; p-values of pairwise comparison between genotype subgroups are ≥ 0.1 and are not reported in the Table.

**eTable 7. Impact of *HIF1A* genotype on NFL levels after treatment start.**

| | ***N*** | ***Years [SD]*** | ***Beta*** | ***95% CI*** | ***P*** |
|---|---|---|---|---|---|
| ***Dimethyl Fumarate*** | 151 | 1.07 [0.12] | -0.38 | -0.71 to -0.043 | **.027** |
| ***Teriflunomide*** | 52 | 0.90 [0.24] | -0.40 | -0.94 to 0.14 | .15 |
| ***Natalizumab*** | 178 | 1.06 [0.28] | -0.16 | -0.45 to 0.13 | .28 |
| ***Fingolimod*** | 130 | 1.05 [0.17] | -0.18 | -0.57 to 0.21 | .36 |

The table shows the results of the mixed-effects linear models assessing the effect of rs1951795 genotype on the change in plasma NFL levels after treatment start. For each drug, the table reports: the number of subjects included (N), the mean time in years (with SD = standard deviation) between the first plasma NFL measurement (obtained immediately before drug start) and the second plasma NFL measurement (obtained after 12 months with a ±6-month window), the regression coefficient from mixed-effect linear model (beta) and it's 95% confidence interval (CI) and p-values (P) from mixed-effects linear models.

### REFERENCES

1. Thompson AJ, Banwell BL, Barkhof F, et al. Diagnosis of multiple sclerosis: 2017 revisions of the McDonald criteria. *Lancet Neurol* 2018;17(2):162–73.
2. the Haplotype Reference Consortium. A reference panel of 64,976 haplotypes for genotype imputation. *Nat Genet* 2016;48(10):1279–83.
3. Ashburner M, Ball CA, Blake JA, et al. Gene Ontology: tool for the unification of biology. *Nat Genet* 2000;25(1):25–9.
4. Kanehisa M. KEGG: Kyoto Encyclopedia of Genes and Genomes. *Nucleic Acids Res* 2000;28(1):27–30.
5. Karolchik D. The UCSC Table Browser data retrieval tool. *Nucleic Acids Res* 2004;32(90001):493D – 496.
6. Chang CC, Chow CC, Tellier LC, Vattikuti S, Purcell SM, Lee JJ. Second-generation PLINK: rising to the challenge of larger and richer datasets. *GigaScience* 2015;4(1):7.
7. Gabriel SB, Schaffner SF, Nguyen H, et al. The Structure of Haplotype Blocks in the Human Genome. *Science* 2002;296(5576):2225–9.
8. Dobin A, Davis CA, Schlesinger F, et al. STAR: ultrafast universal RNA-seq aligner. *Bioinformatics* 2013;29(1):15–21.
9. Bolger AM, Lohse M, Usadel B. Trimmomatic: a flexible trimmer for Illumina sequence data. *Bioinformatics* 2014;30(15):2114–20.
10. Liao Y, Smyth GK, Shi W. featureCounts: an efficient general purpose program for assigning sequence reads to genomic features. *Bioinformatics* 2014;30(7):923–30.
11. Ewels P, Magnusson M, Lundin S, Käller M. MultiQC: summarize analysis results for multiple tools and samples in a single report. *Bioinformatics* 2016;32(19):3047–8.
12. Valverde S, Cabezas M, Roura E, et al. Improving automated multiple sclerosis lesion segmentation with a cascaded 3D convolutional neural network approach. *NeuroImage* 2017;155:159–68.
13. Schmidt P, Gaser C, Arsic M, et al. An automated tool for detection of FLAIR-hyperintense white-matter lesions in Multiple Sclerosis. *NeuroImage* 2012;59(4):3774–83.
14. Kuhlmann T, Ludwin S, Prat A, Antel J, Brück W, Lassmann H. An updated histological classification system for multiple sclerosis lesions. *Acta Neuropathol* (Berl) 2017;133(1):13–24.
15. Bankhead P, Loughrey MB, Fernández JA, et al. QuPath: Open source software for digital pathology image analysis. *Sci Rep* 2017;7(1):16878.
16. DeLuca GC. The contribution of demyelination to axonal loss in multiple sclerosis. *Brain* 2006;129(6):1507–16.
17. Diaz-Sanchez M, Williams K, DeLuca GC, Esiri MM. Protein co-expression with axonal injury in multiple sclerosis plaques. *Acta Neuropathol* (Berl) 2006;111(4):289–99.
18. Hillert J, Stawiarz L. The Swedish MS registry – clinical support tool and scientific resource. *Acta Neurol Scand* 2015;132(S199):11–9.
19. Manouchehrinia A, Stridh P, Khademi M, et al. Plasma neurofilament light levels are associated with risk of disability in multiple sclerosis. *Neurology* 2020;94(23):e2457–67.
20. Huang J, Khademi M, Fugger L, et al. Inflammation-related plasma and CSF biomarkers for multiple sclerosis. *Proc Natl Acad Sci* 2020;117(23):12952–60.
21. McCaw ZR, Lane JM, Saxena R, Redline S, Lin X. Operating characteristics of the rank-based inverse normal transformation for quantitative trait analysis in genome-wide association studies. *Biometrics* 2020;76(4):1262–72.
22. Benkert P, Meier S, Schaedelin S, et al. Serum neurofilament light chain for individual prognostication of disease activity in people with multiple sclerosis: a retrospective modelling and validation study. *Lancet Neurol* 2022;21(3):246–57.
23. Havrdova E, Galetta S, Stefoski D, Comi G. Freedom from disease activity in multiple sclerosis. *Neurology* 2010;74(Issue 17, Supplement 3):S3–7.
